# Long-term health-related quality of life in non-hospitalised COVID-19 cases with confirmed SARS-CoV-2 infection in England: Longitudinal analysis and cross-sectional comparison with controls

**DOI:** 10.1101/2021.10.22.21264701

**Authors:** Frank Sandmann, Elise Tessier, Joanne Lacy, Meaghan Kall, Edwin Van Leeuwen, Andre Charlett, Rosalind M Eggo, Gavin Dabrera, W. John Edmunds, Mary Ramsay, Helen Campbell, Gayatri Amirthalingam, Mark Jit

**Affiliations:** Statistics, Modelling and Economics Department, UK Health Security Agency, 61 Colindale Ave, London NW9 5EQ, UK; Department of Infectious Disease Epidemiology, London School of Hygiene and Tropical Medicine, Keppel St, London WC1E 7HT, UK; Immunisation Division, UK Health Security Agency, 61 Colindale Ave, London NW9 5EQ, UK; COVID-19 National Epidemiology Cell, UK Health Security Agency, Wellington House, 133-155 Waterloo Rd, London SE1 8UG, UK

**Keywords:** SARS-CoV-2, COVID-19, long COVID, health-related quality of life, QALYs

## Abstract

**Background:** This study measured the long-term health-related quality of life of non-hospitalised COVID-19 cases with PCR-confirmed SARS-CoV-2(+) infection using the recommended instrument in England (the EQ-5D).

**Methods:** Prospective cohort study of SARS-CoV-2(+) cases aged 12-85 years and followed up for six months from 01 December 2020, with cross-sectional comparison to SARS-CoV-2(-) controls. Main outcomes were loss of quality-adjusted life days (QALDs); physical symptoms; and COVID-19-related private expenditures. We analysed results using multivariable regressions with post-hoc weighting by age and sex, and conditional logistic regressions for the association of each symptom and EQ-5D limitation on cases and controls.

**Results:** Of 548 cases (mean age 41.1 years; 61.5% female), 16.8% reported physical symptoms at month 6 (most frequently extreme tiredness, headache, loss of taste and/or smell, and shortness of breath). Cases reported more limitations with doing usual activities than controls. Almost half of cases spent a mean of £18.1 on non-prescription drugs (median: £10.0), and 52.7% missed work or school for a mean of 12 days (median: 10). On average, all cases lost 15.9 (95%-CI: 12.1, 19.7) QALDs, while those reporting symptoms at month 6 lost 34.1 (29.0, 39.2) QALDs. Losses also increased with older age. Cumulatively, the health loss from morbidity contributes at least 21% of the total COVID-19-related disease burden in England.

**Conclusions:** One in 6 cases report ongoing symptoms at 6 months, and 10% report prolonged loss of function compared to pre-COVID-19 baselines. A marked health burden was observed among older COVID-19 cases and those with persistent physical symptoms.

**summary:** Losses of health-related quality of life in non-hospitalised COVID-19 cases increase by age and for cases with symptoms after 6 months. At a population level, at least 21% of the total COVID-19-related disease burden in England is attributable to morbidity.

## Introduction

Severe acute respiratory syndrome coronavirus 2 (SARS-CoV-2) causing coronavirus disease 2019 (COVID-19) is associated with substantial morbidity, mortality, and costs to healthcare and society. There is a growing body of evidence on persistent physical symptoms in some individuals who recover from COVID-19 (sometimes called “long COVID”), with a reported prevalence of 20-30% after one month and at least 10% after three months [1]. Cases with post-acute COVID-19 syndrome also report a worsened health-related quality of life (HRQoL) [2].

Robust estimates of the impact of COVID-19 in terms of quality-adjusted life years (QALYs) lost are needed to inform policy. These values are essential for costs-per-QALY estimates to evaluate health technologies such as non-pharmaceutical interventions, therapeutics, diagnostics, and vaccines. For instance, recommendations by the National Institute of Health and Care Excellence (NICE) and the Joint Committee on Vaccination and Immunisation (JCVI) in the UK require such estimates [3, 4]. The disease-generic EQ-5D is a validated, patient-reported instrument and is the recommended method to estimate QALY losses in England [3, 4] and many other countries [5]. The importance of COVID-19-specific HRQoL estimates is growing [6], particularly as countries continue facing the impact of post-acute COVID-19 syndrome [1].

Therefore, we aimed to assess the HRQoL impact of SARS-CoV-2 infection and COVID-19-like symptoms over 6 months in England using the EQ-5D. The primary aim was to provide COVID-19-specific QALY loss estimates of non-hospitalised confirmed cases. We also explored the physical symptom status and any private expenditures due to COVID-19 of cases. Lastly, we compared the SARS-CoV-2(+) cases cross-sectionally at month 6 to SARS-CoV-2(-) controls.

## Methods

### Study design and patient population (SARS-CoV-2 positive cases)

We report on a prospective observational study of individuals with non-fatal SARS-CoV-2 infection and COVID-19 symptoms in England, with a cross-sectional comparison to controls matched by region at month 6 (see next section). Infections with SARS-CoV-2 were laboratory-confirmed by real-time polymerase chain reaction (RT-PCR) in November 2020. Participants aged 12 to 85 years inclusive were randomly selected from the Second Generation Surveillance System (SGSS; the routine laboratory reporting system in England [7]) to be proportionately representative by geographical region in England (East Midlands, East of England, London, North East, North West, South East, South West, West Midlands, Yorkshire and The Humber) from all those who requested a test through community testing with a specimen date of 26-27 November 2020. At the time, the lineage of cases in England were predominantly formed by wild type and increasingly the Alpha variant (S-gene negative B.1.1.7) [8-10]. Laboratory confirmed cases were invited by email and post to this study on or shortly after December 01, 2020. Other than non-fatal outcome, non-hospitalised, PCR-positive, and aged between 12 and 85 years there were no formal recruitment restrictions for cases. The study is an interim evaluation of ongoing surveillance, with reporting here on the first 6 months post-infection

Surveys were sent out electronically using Snap Surveys (www.snapsurveys.com), and the initial survey was also sent by post to all participants to increase response rates. Subsequent surveys were sent out via the method that the participant used to respond to the first survey.

The initial survey enquired about baseline demographics (age, sex, pregnancy, ethnicity, and comorbidities), physical symptoms (in the first 7 days of illness given the potential delay in symptom onset, getting PCR tested, and receiving the survey, and at the time for filling out the survey), resource use and personal expenditures due to COVID-19 not falling on the healthcare system (like absence days from work or school, time used to care for others or being cared for, and costs for medication and other help due to the COVID-19 illness episode), and the HRQoL impact as measured through the adult version of the EQ-5D-5L, which collects information about 5 dimensions (mobility, self-care, usual activities, pain/discomfort, anxiety/depression) on 5 levels (no problems, slight problems, moderate problems, severe problems, extreme problems). The EQ-5D is validated to be used with the visual analogue scale (VAS), which is a rating scale from 0 to 100 that indicates the worst to best imaginable health, respectively. Given that cases were recruited after having tested positive for SARS-CoV-2, we used the EQ-5D three times in the initial survey for participants to self-report their health at three time points: 1) health on the day they filled in the survey, 2) pre-COVID-19 baseline health, and 3) health on the worst day of COVID-19.

After the initial survey, participants received short follow-up surveys sent out at week 2, week 4, week 6, week 8, week 12 and month 6. Before sending out each survey we cross-checked mortality of respondents to minimise emotional distress in bereaved family members (n < 5; precise number suppressed to prevent data disclosure issues). Respondents were also reminded that they could opt out of the study at the end of each survey.

During the study design phase in spring/summer 2020, we estimated the sample size required with 300-350 cases to detect a difference with 80% power at a significance level of 5% and assuming a difference of half a quality-adjusted life-day (QALD) lost by laboratory-confirmed cases. This assumption was informed by similar studies on the quality of life during the H1N1pdm2009 influenza pandemic [11, 12]. To account for an anticipated response rate of 20%, we invited 1,500 laboratory-confirmed cases to participate. We received a response to the initial survey of 1,500 from 566 individuals (response rate of 37.7%). After removal of 11 delayed responses that overlapped with the next follow-up survey and seven cases who received hospital care, we analysed the responses of 548 cases (with 157 cases still responding at month 6).

### SARS-CoV-2 negative controls

At the time of the follow-up survey in June 2021, we also recruited a negative control group of individuals who had a negative SARS-CoV-2 test on 26-27 November 2020 and had no subsequent positive SARS-CoV-2 test. The controls were identified in SGSS as individuals with a negative PCR test with a sample date on 26 or 27 November 2020. They were frequency-matched to the invited cases by the nine geographical regions in England to reflect the spatial distribution of the SARS-CoV-2 positive cases. To reduce administrative burden we invited controls by email, and randomly selected individuals to achieve a match of three controls per case. After excluding individuals without email addresses, and those reported to have died, the online survey was sent to 4,908 controls. The survey asked about the symptom status of controls for other illnesses between March and June 2021. The EQ-5D-5L was used to ask about their health on the day of the survey. Based on the received responses of 750 individuals (15.3%), we limited the controls to the same age range as the cases. We also excluded 15 individuals who self-identified as having tested SARS-CoV-2 positive e.g. as part of clinical trials or abroad, leaving a total of 651 controls for the cross-sectional analysis at month 6.

### Statistical analysis

We summarised the baseline characteristics of respondents in terms of demography. To evaluate representativeness, we compared these with (1) the limited demographic information we had for non-respondents, and (2) the population in England. For respondents, we further analysed the symptom status, COVID-19-related resource use and private expenditures, and the health-related quality of life as measured on the 5 domains of the EQ-5D and the VAS. Changes of the EQ-5D responses over time were also described as compared to the pre-COVID baseline [13].

To calculate QALYs lost due to COVID-19, we first mapped EQ-5D results to the utility value set of the EQ-5D-5L and then cross-walked results to the EQ-5D-3L in England [14-16]. In sensitivity analysis, we used different international value sets [17-20] (see Appendix for details). We then calculated individual patient-level QALYs using the area-under-the-curve (AUC) of the utility values and the calendar dates between completed surveys. We then estimated QALY losses due to COVID-19 per patient based on dis-utilities, i.e. the difference of a utility value indicating perfect health (of 1.0). Separately, we also calculated QALY losses of recovered participants based on only the pre-COVID baseline, the worst day of illness, and the subsequent value once recovered per individual (at any timepoint of the follow up surveys).

We analysed the total individual patient-level QALY loss over 6 months as the dependent variable in linear regression models with bias-corrected and accelerated confidence intervals based on the nonparametric bootstrap [21]. The first regression was a marginal model of cases reporting being symptomatic at month 6 (coded as symptomatic = 1, asymptomatic = 0) against the QALY loss. The second regression model explored adjusting for the impact of age, sex, pregnancy status, ethnicity, the presence of comorbidities, having been told to shield in the early phase of the pandemic (“shielding”) [22], reporting being symptomatic at month 6, the household size, geographical region, the mode of survey submission (online or postal), the reason for being tested, the strain variant (Wildtype or Alpha), being vaccinated against seasonal influenza in 2020-2021, being fully vaccinated against COVID-19, and the pre-COVID-19 baseline utility [23]. Variables with *p* < 0.1 in univariate analyses were entered into the multivariable analyses, and tested for multicollinearity. We repeated this estimation for each of the 6 different EQ-5D value sets as dependent variable. No responses were imputed given the large proportion of missing values [24].

We then calculated the COVID-19-related burden at the population-level using our estimated QALY losses per case by age, which we compared to the QALY losses per death by age [25] (considering the 6.4 million confirmed cases and 118,858 COVID-19-related deaths in England by September 19, 2021; https://coronavirus.data.gov.uk/). We also used post-hoc weighting by age group and sex to account for the imbalances in our sample versus the population in England.

For ease of presentation, we converted the results in the main text to quality-adjusted life days (QALDs) by multiplying the QALYs with a value of 365.25 days.

For the cross-sectional analysis at month 6, we separately analysed the differences in physical symptoms and health utility index values between the SARS-CoV-2(+) cases and the SARS-CoV-2(-) controls. We used conditional logistic regression models based on the matched geographical regions to investigate the association of each symptom and EQ-5D dimension with the persistence of symptoms by month 6 (with the EQ-5D dimensions coded as 0 = no limitation, 1 = any limitation). For the continuous EQ-5D health utility index values at month 6, we used multivariable linear regression models after obtaining a singular fit that suggests no heterogeneity between regions when using a mixed-effects regression model with regions as random effect. Therefore, we followed a similar approach as described above (without the covariates of the mode of survey submission, reason for being tested, strain variant, and baseline utility that were either a constant or not collected for the controls). We included an additional indicator variable to distinguish cases from controls. For the analysis looking at each symptom we used the data of both cases and controls at month 6, while for the analysis looking at the impact on the EQ-5D dimensions we used the data of cases and controls as well as stratified for cases alone, which allows comparing SARS-CoV-2(+) cases with persistent symptoms versus SARS-CoV-2(+) cases who recovered by month 6.

Given that the respondents reported on results for fewer than 12 months we did not apply discount rates [3, 4].

All analyses were conducted in R (R Core Team, Vienna, Austria).

### Ethics

Ethical approval for this study was not required as the collection of information on patients’ quality of life is part of the statutory tasks and routine surveillance activities of Public Health England and its legal successor, the UK Health Security Agency.

## Results

### Participant characteristics and descriptive statistics

The mean age of cases was 41.1 years at baseline (95% uncertainty interval, 95%-UI: 13.0-74.0), and 61.5% identified as female (81.6%; Table 1). At month 6, the mean age of responding cases was 49.4 years (13.9-78.1), and 61.8% identified as female. This compares with the controls who had a mean age of 45.4 years (16.0-72.8) and 70.7% identifying as female. Further demographic details of the participants are summarised in Table 1. Our respondents over-represented middle-aged adults, women, people of white ethnicity and individuals from London than the non-respondents and the population in England (Supplementary Figures 1 to 4). Post-stratification weights by age and sex were applied to partially correct for these differences and ensure the sample is more representative of cases in England at that time

**Table 1.**
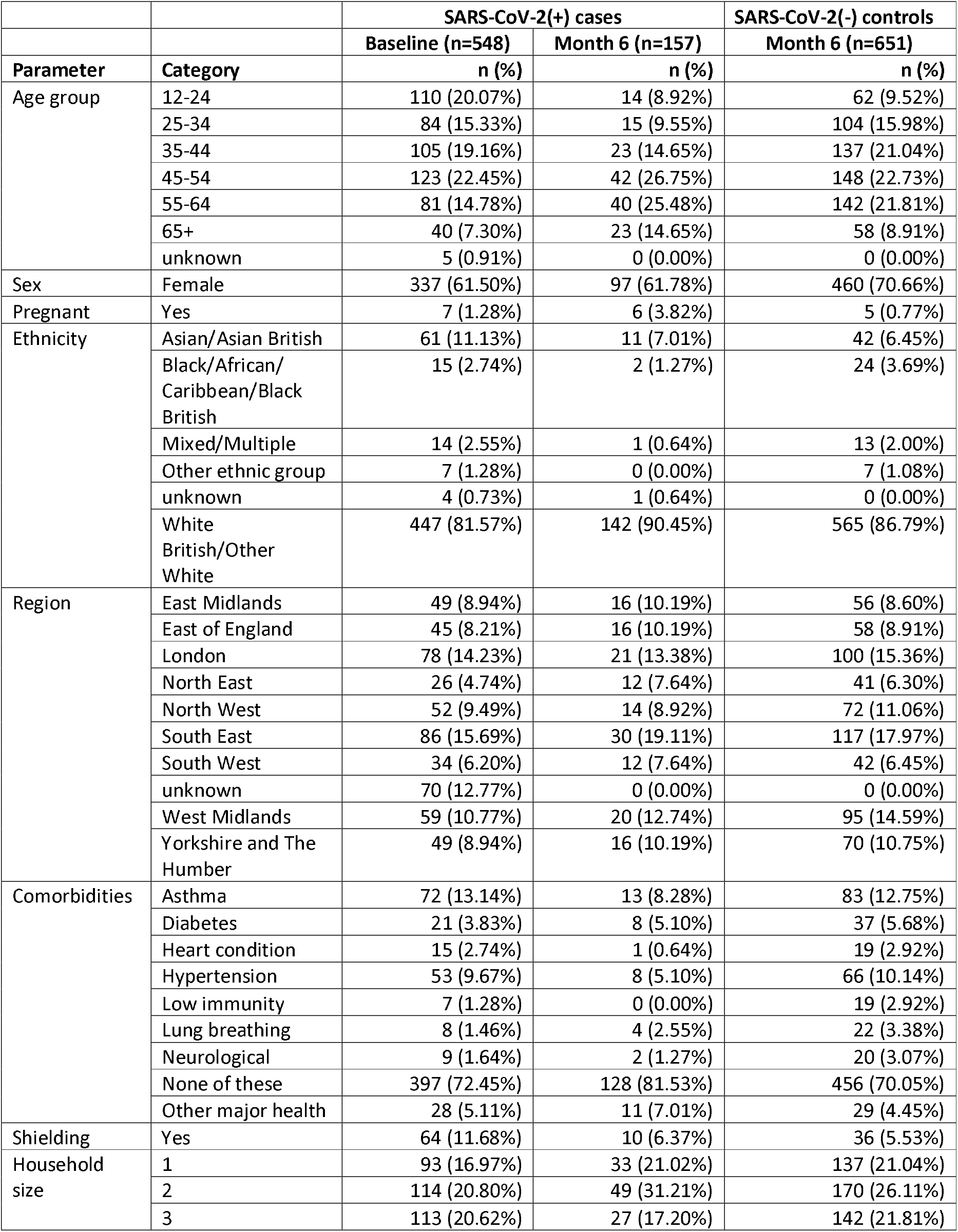
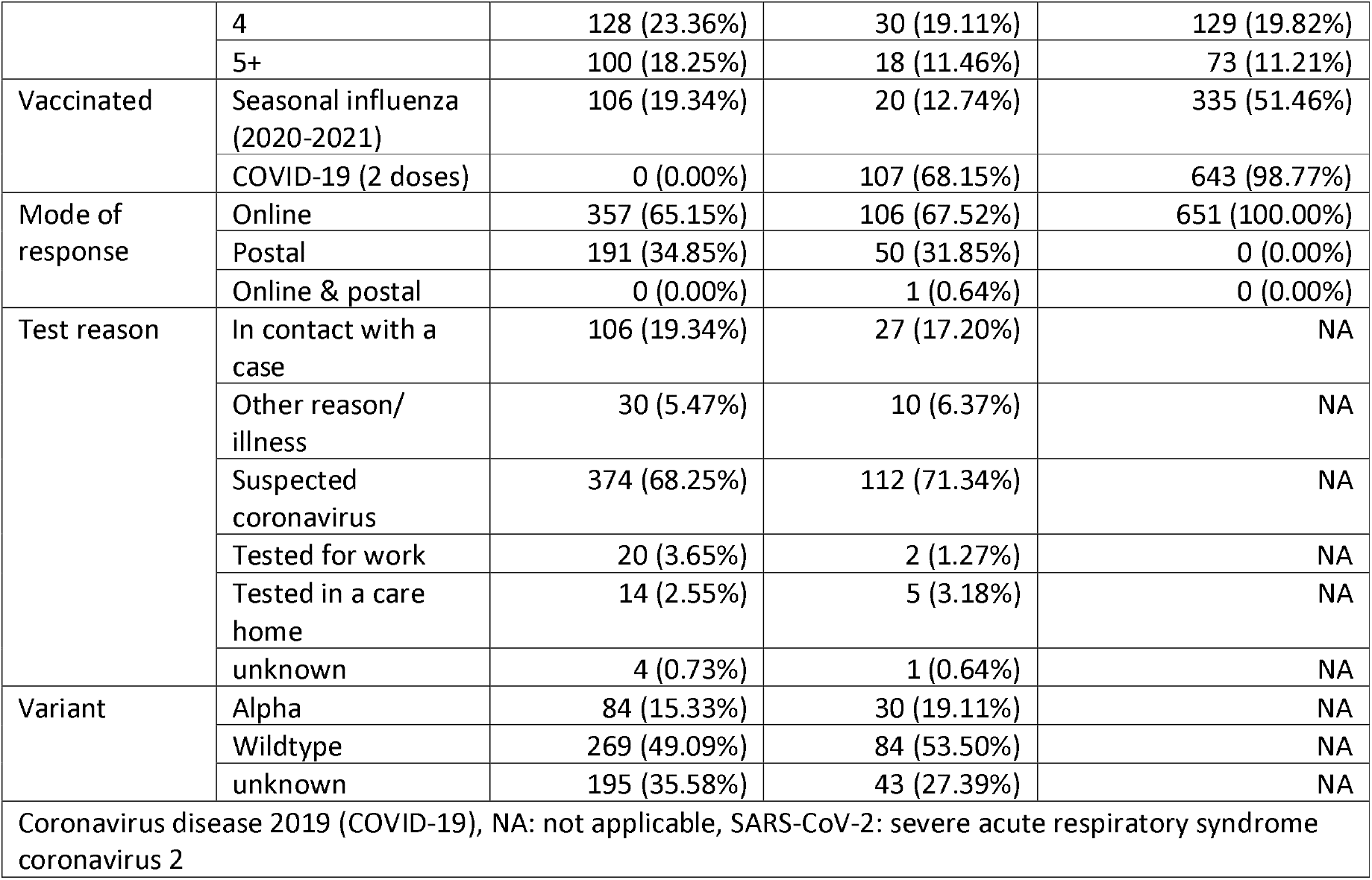
Demographic characteristics and summary of the SARS-CoV-2(+) cases at baseline, and of the SARS-CoV-2(+) cases and SARS-CoV-2(-) controls at month 6.

### Physical symptoms, healthcare resource use and private expenditures

Over 95% of cases reported any symptom in the first 7 days of being tested or onset of being ill, which decreased to a weighted mean of 35.7% (183/548) at week 4, 20.4% (109/548) at week 12 and 19.7% (92/548) by month 6. More than 50% reported symptoms of headache, extreme tiredness/lack of energy, loss of taste and/or loss of smell, and muscle aches (Figure 1). At month 6, the most frequently reported symptoms were extreme tiredness, headache, loss of taste and smell, and shortness of breath. Unweighted estimates were very similar to the weighted estimates (Supplementary Figure 5).

**Figure 1:**
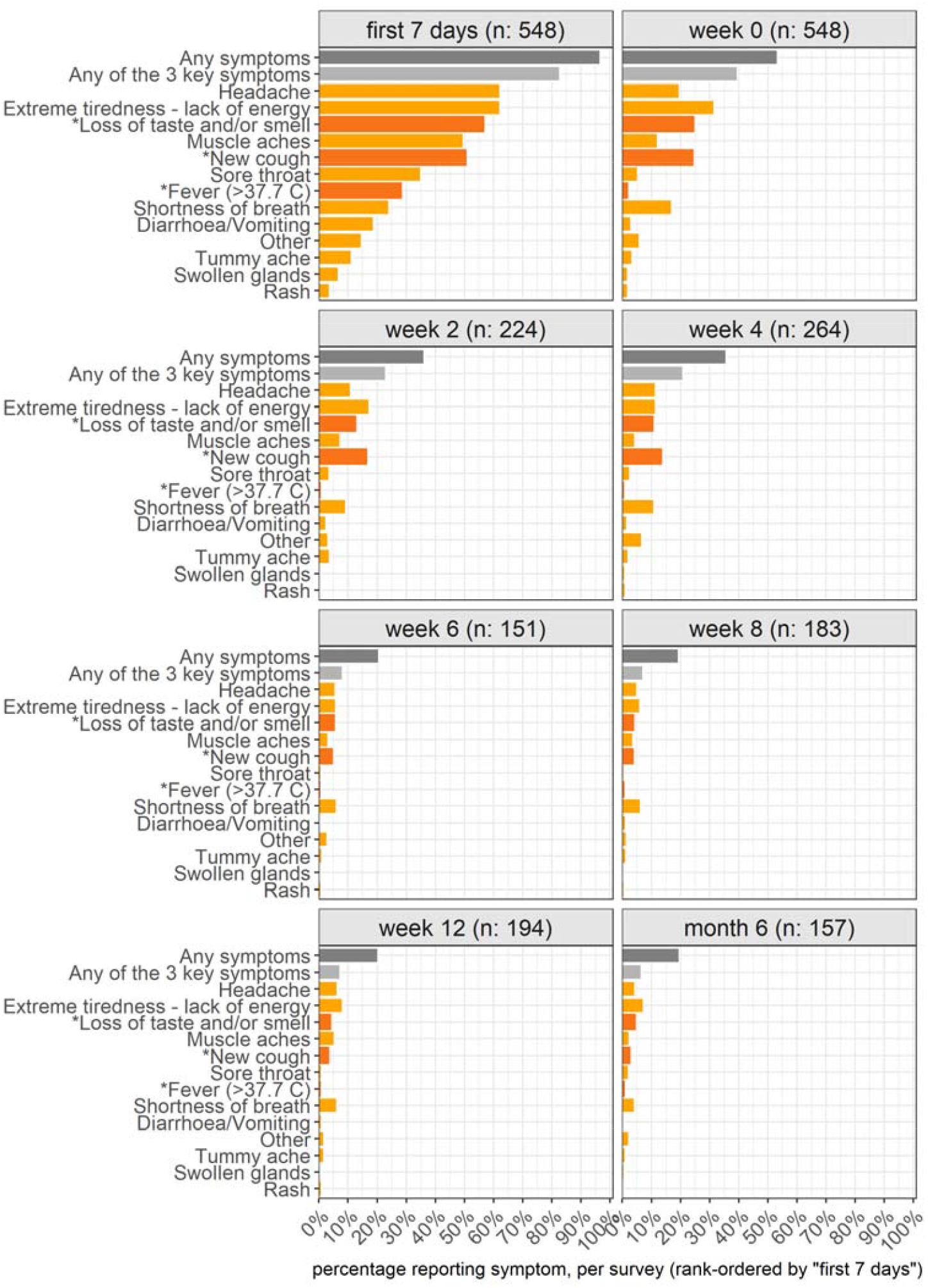
Physical symptoms reported in each survey by the SARS-CoV-2(+) cases over 6 months weighted by age and sex of the population in England. Symptoms that have been seen as key indicators in the UK for members of the public to request a PCR test for COVID-19 are indicated in a darker colour and starred (i.e., fever of 38.8 C or higher, a new cough, or loss of taste and/or smell). The panels are rank-ordered by the proportion of symptoms in the first 7 days (top-left panel). Given the potential delay in symptom onset, getting PCR tested, and receiving the initial survey, the first survey enquired about the symptom status of cases within the first 7 days of feeling ill as well as if still feeling unwell today (week 0).

In terms of healthcare resource use (excluding hospital care), cases most frequently reported GP visits (15.5%; Table 2). Furthermore, 45.3% of cases reported taking non-prescription medicines such as pain killers to manage their COVID-19 illness, with private expenditures of a mean of £18.07 (median: £10.00, range: £0.50, £200.00). 38.0% of cases reported being cared for by someone else and/or caring for someone else with COVID-19. In both situations, the duration of caring was for a mean of 11 days (Table 2). Another 52.7% of cases missed work or school due to their COVID-19 episode, for an estimated total of a mean of 12 days (which may partly reflect infection control legislation that required 10 days of self-isolation). 20.8% of cases reported receiving other paid help, with mean expenditures of £61.35 (median: £25.00, range: £2.00, £1,000.00).

**Table 2.**
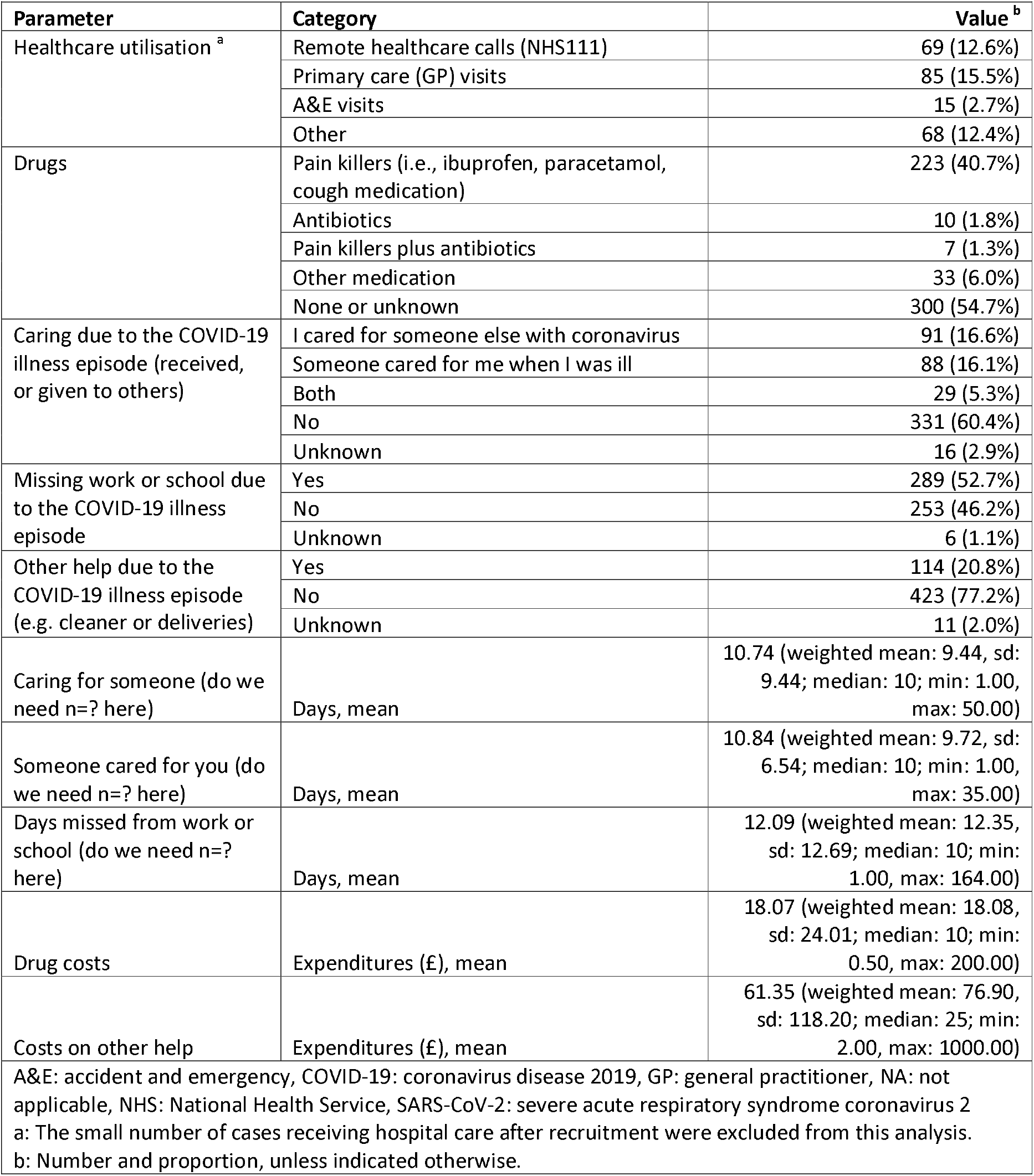
Frequencies of healthcare resource use, care taking, work and school absences, and receiving other help due to the COVID-19 illness episode, and personal costs of the SARS-CoV-2(+) cases over 6 months.

|  |  |  |
| --- | --- | --- |
| Parameter | Category | Value <sup>b</sup> |
| Healthcare utilisation <sup>a</sup> | Remote healthcare calls (NHS111) | 69 (12.6%) |
|  | Primary care (GP) visits | 85 (15.5%) |
|  | A&E visits | 15 (2.7%) |
|  | Other | 68 (12.4%) |
| Drugs | Pain killers (i.e., ibuprofen, paracetamol, cough medication) | 223 (40.7%) |
|  | Antibiotics | 10 (1.8%) |
|  | Pain killers plus antibiotics | 7 (1.3%) |
|  | Other medication | 33 (6.0%) |
|  | None or unknown | 300 (54.7%) |
| Caring due to the COVID-19 illness episode (received, or given to others) | I cared for someone else with coronavirus | 91 (16.6%) |
|  | Someone cared for me when I was ill | 88 (16.1%) |
|  | Both | 29 (5.3%) |
|  | No | 331 (60.4%) |
|  | Unknown | 16 (2.9%) |
| Missing work or school due to the COVID-19 illness episode | Yes | 289 (52.7%) |
|  | No | 253 (46.2%) |
|  | Unknown | 6 (1.1%) |
| Other help due to the COVID-19 illness episode (e.g. cleaner or deliveries) | Yes | 114 (20.8%) |
|  | No | 423 (77.2%) |
|  | Unknown | 11 (2.0%) |
| Caring for someone (do we need n=? here) | Days, mean | 10.74 (weighted mean: 9.44, sd: 9.44; median: 10; min: 1.00, max: 50.00) |
| Someone cared for you (do we need n=? here) | Days, mean | 10.84 (weighted mean: 9.72, sd: 6.54; median: 10; min: 1.00, max: 35.00) |
| Days missed from work or school (do we need n=? here) | Days, mean | 12.09 (weighted mean: 12.35, sd: 12.69; median: 10; min: 1.00, max: 164.00) |
| Drug costs | Expenditures (£), mean | 18.07 (weighted mean: 18.08, sd: 24.01; median: 10; min: 0.50, max: 200.00) |
| Costs on other help | Expenditures (£), mean | 61.35 (weighted mean: 76.90, sd: 118.20; median: 25; min: 2.00, max: 1000.00) |
| A&E: accident and emergency, COVID-19: coronavirus disease 2019, GP: general practitioner, NA: not applicable, NHS: National Health Service, SARS-CoV-2: severe acute respiratory syndrome coronavirus 2<br>a: The small number of cases receiving hospital care after recruitment were excluded from this analysis.<br>b: Number and proportion, unless indicated otherwise. |  |  |

### Health-related quality of life (EQ-5D), index utilities, and QALY losses due to COVID-19

The proportion of cases reporting any problems on the EQ-5D was lowest before the COVID-19 episode and less than 10% for the dimensions of self-care, mobility, and usual activities, less than 20% for pain / discomfort and less than 30% for anxiety/depression (Figure 2a). In comparison, on the worst day of the COVID-19 episode more than 80% of cases reported experiencing pain or discomfort. By week,2, the proportion who reported problems with self-care was restored to pre-COVID-19 baseline levels, while problems on the other dimensions returned to pre-COVID-19 baselines around week 8. Notably, slight-to-moderate anxiety / depression were observed most frequently at the pre-COVID-19 baseline and again after week 2 through month 6 (Figure 2a, Supplementary Figure 6).

**Figure 2.**
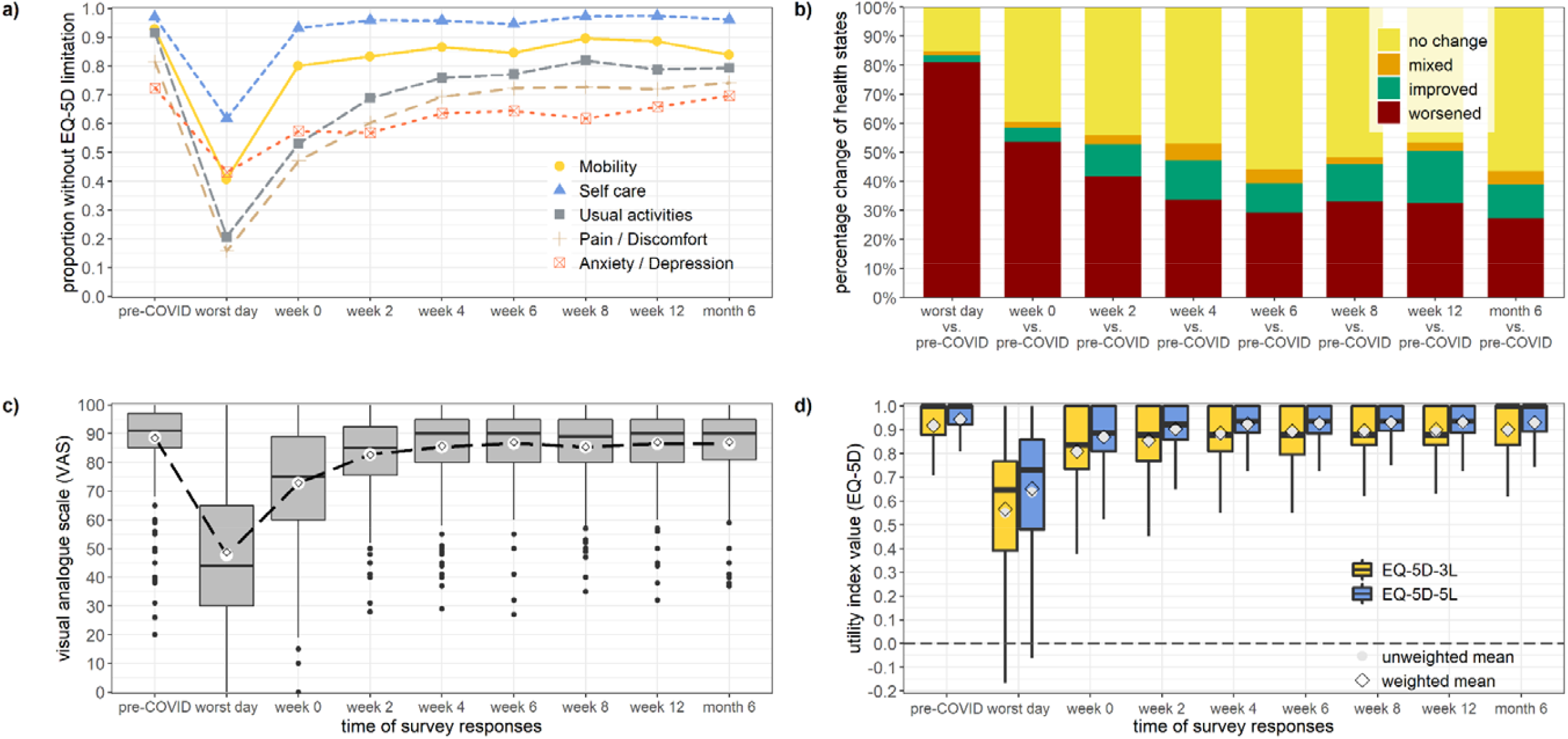
Results on the EQ-5D by the SARS-CoV-2(+) cases in each survey. Proportion of cases without any limitations reported per survey (panel a); percentage change of health states per survey (panel b); mean value of the visual analogue scale (panel c); EQ-5D utility index values using the UK value sets (panel d).

Compared to the pre-COVID-19 baseline, 81% of cases reported a worse health state on the worst day of their illness, which decreased to 27% by month 6 (Figure 2b). However, 69.3% of cases reported no issues at their last recorded observation.

Using the VAS and the EQ-5D index utility values, responses followed a similar trend to the responses on the individual domains in dropping off sharply on the worst day of illness, and recovering by week 4-6 (Figure 2). Index utilities mapped to the EQ-5D-3L were slightly lower than those of the EQ-5D-5L for the UK, and at similar levels to the other international value sets (Supplementary Figure 7).

With a mean follow-up duration of 77.9 days (weighted mean: 84.5 days), the unadjusted health loss due to COVID-19 ranged between 11.3 and 16.4 QALDs with the EQ-5D value sets of different countries (Supplementary Table 1). The unadjusted health loss for just the recovered individuals was lower between 8.8 and 12.1 QALDs (Supplementary Table 1).

The adjusted health loss of the COVID-19 cases were 15.9 (95%-CI: 12.1, 19.7) QALDs with the UK value set for the EQ-5D-3L (Table 3), which also resulted in the highest losses among value sets (Supplementary Table 2). Notably, the health losses were higher for cases who returned surveys by post than online (Supplementary Figure 8), despite being of similar age (47.9 vs 43.0 years) and sex (60.2% female vs 63.8%, respectively). Generally, the health loss of cases increased by age from 9.2 (6.8, 11.5) QALDs in ages 12-24 years to 24.3 (17.5, 31.1) QALDs in ages 65+ years, and it was higher for cases who reported symptoms at month 6 (34.1, 95%-CI: 29.0, 39.2) QALDs versus cases who did not report symptoms at month 6 (10.9, 95%-CI: 8.4, 13.4) QALDs; see Figure 3a, Table 3, and Supplementary Table 3. Cumulatively, the COVID-19-related QALY loss from morbidity contributes 21.0% of the total QALY loss attributed to COVID-19 morbidity and mortality in England by mid-September 2021, and it was higher in age groups younger than 45-54 years than the losses from COVID-19-related mortality (Figure 3b).

**Table 3:** Adjusted QALD losses of the SARS-CoV-2(+) cases, and by SARS-CoV-2(+) cases who report symptoms at month 6.

| Value set (EQ-5D) | Group | Unweighted results |  |  | Weighted results |  |  |
| --- | --- | --- | --- | --- | --- | --- | --- |
|  |  | Estimate | Lower 95%-CI | Upper 95%-CI | Estimate | Lower 95%-CI | Upper 95%-CI |
| UK (3L) | All cases | 15.0 | 14.0 | 16.0 | 15.9 | 12.1 | 19.7 |
| UK (5L) |  | 12.0 | 11.2 | 12.7 | 12.7 | 9.9 | 15.6 |
| UK (3L) | Cases without symptoms at month 6 | 10.6 | 9.9 | 11.2 | 11.0 | 8.5 | 13.6 |
| UK (5L) |  | 8.8 | 8.3 | 9.3 | 9.2 | 7.2 | 11.1 |
| UK (3L) | Cases with symptoms at month 6 | 36.9 | 35.3 | 38.5 | 36.9 | 29.2 | 39.2 |
| UK (5L) |  | 27.7 | 26.5 | 28.9 | 27.6 | 21.9 | 29.4 |

CI: confidence interval, QALD: quality-adjusted life day, QALY: quality-adjusted life year, UK: United Kingdom.
Results are based on the final regression models of the QALY loss per EQ-5D value set and adjusted for baseline utility, reporting symptoms at month 6, age, pregnancy, household size, mode of response, having received the influenza vaccine in 2020-2021, and having received 2 COVID-19 vaccine doses. We post-hoc weighted results based on age and sex of the population in England.

**Figure 3.**
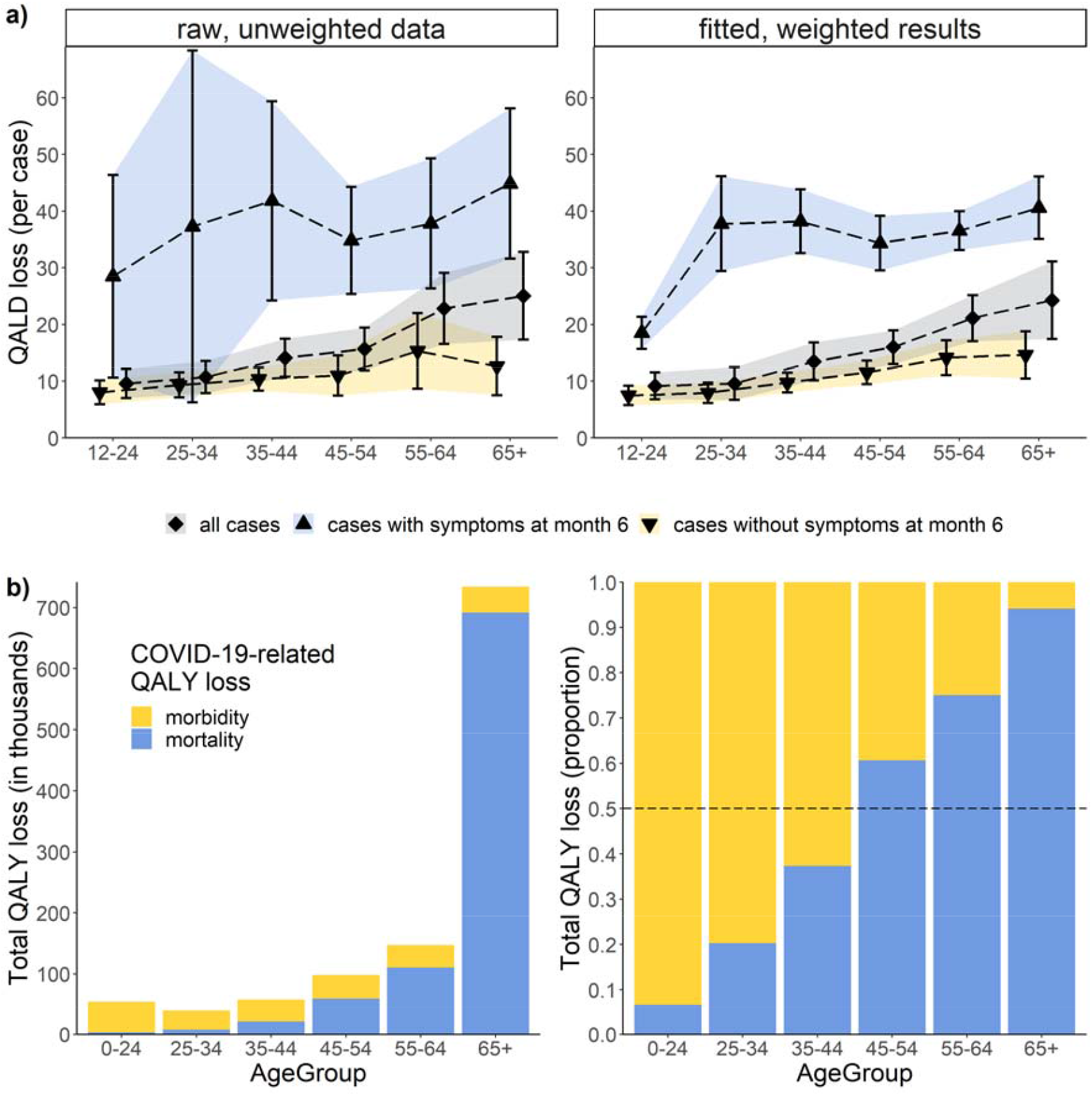
COVID-19-related quality-adjusted life day (QALD) losses per case by age group and presence of reported symptoms at month 6 (panel a ; raw data versus fitted results that were post-hoc weighted by sex), and the total COVID-19-related quality-adjusted life years (QALY) loss of cases versus deaths by age group in England at population-level (panel b). QALDs are equivalent to the number of QALYs^*^365.25 days.

### Cross-sectional analysis of SARS-CoV-2(+) cases and SARS-CoV-2(-) controls

Cases were more likely to report physical symptoms than controls, particularly extreme tiredness, headache, loss of taste and/or smell, shortness of breath, and cough (Figure 3). Cases with persistent symptoms also had more difficulties with muscle aches and other symptoms than controls or recovered cases (Figure 4). In separate EQ-5D dimensions, cases more frequently reported problems with doing usual activities than controls, while controls more frequently reported problems with pain / discomfort and anxiety / depression than cases but differences were not significant (Figure 5). The adjusted utility index values improved for cases without symptoms by month 6 but were worse for symptomatic individuals, irrespective of using the EQ-5D-3L (Table 4) or the EQ-5D-5L (Supplementary Table 4).

**Figure 4.**
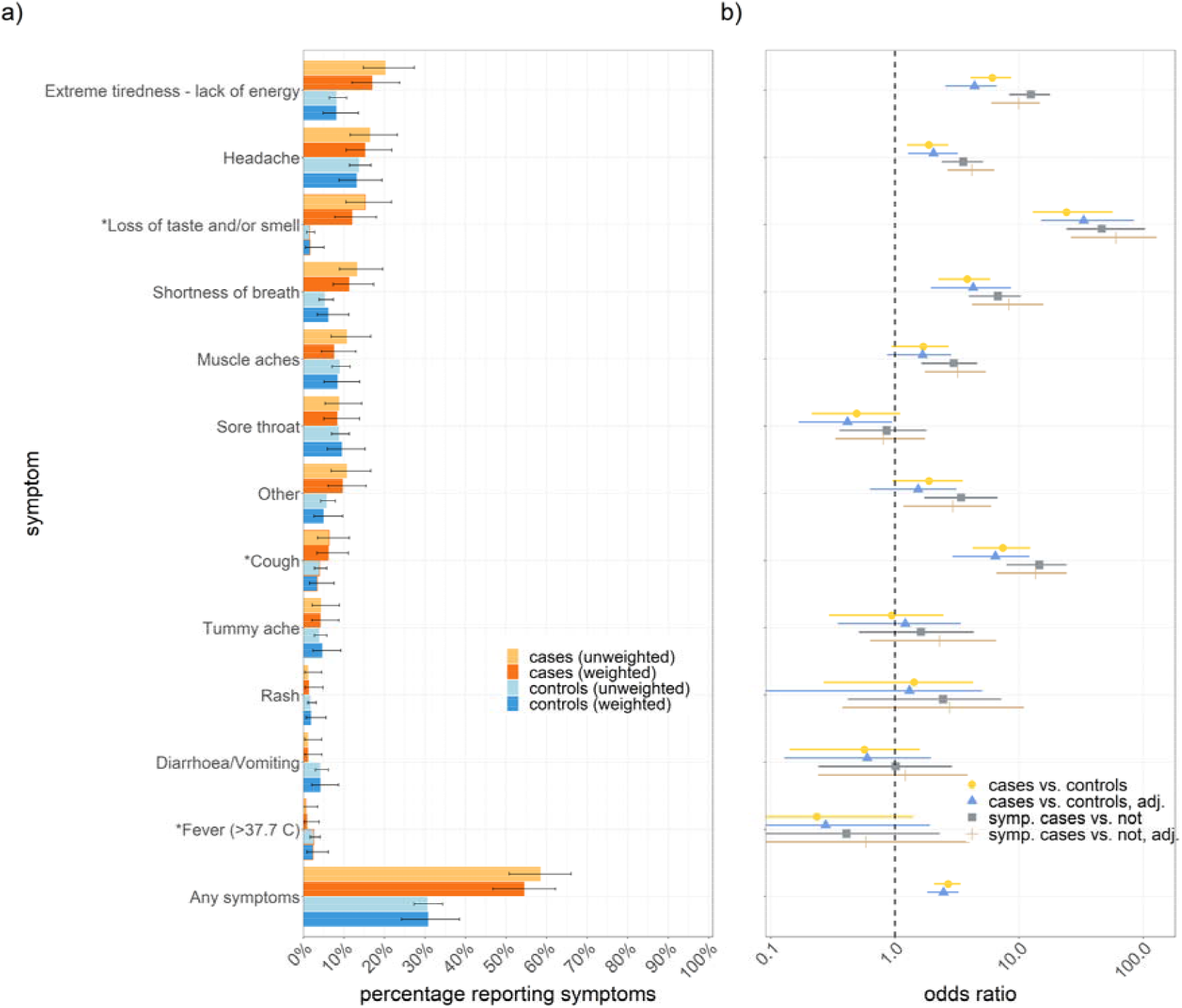
Comparison of the physical symptoms reported by the SARS-CoV-2(+) cases and SARS-CoV-2(-) controls at month 6 (panel a), and the association between cases (and cases with persistent symptoms) and physical symptoms (panel b). Panel a) shows the unadjusted results with 95% binomial confidence intervals (unweighted, and weighted by age and sex), and panels b) shows the conditional logistic regression results.

**Figure 5.**
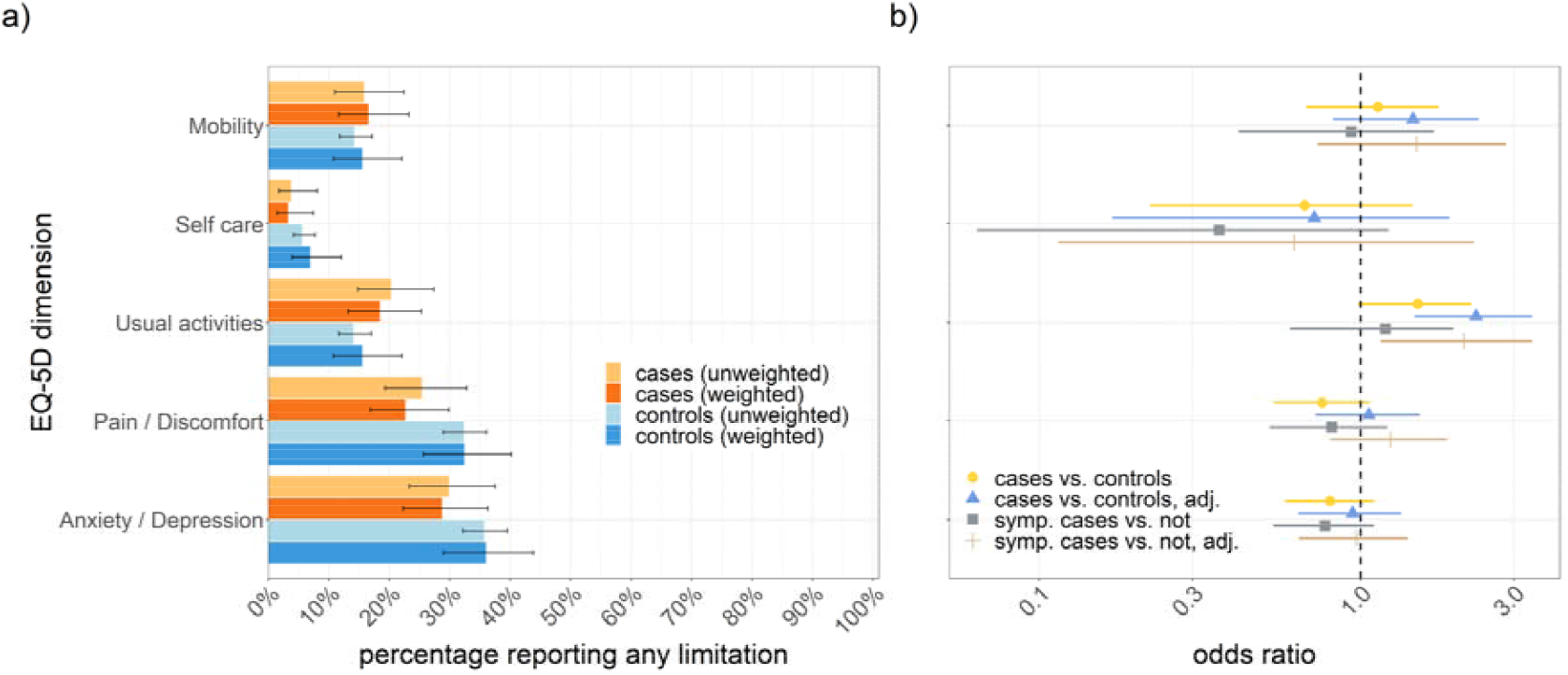
Comparison of the limitations per EQ-5D dimension reported by the SARS-CoV-2(+) cases and SARS-CoV-2(-) controls at month 6 (panel a), and the association between cases (and cases with persistent symptoms) and limitation per EQ-5D dimension (panel b). Panel a) shows the unadjusted results with 95% binomial confidence intervals (unweighted, and weighted by age and sex), and panels b) shows the conditional logistic regression results.

**Table 4.** Multivariable linear regression models of the EQ-5D-3L utility index values of the SARS-CoV-2(+) cases and SARS-CoV-2(-) controls at month 6.

| Parameter | Category | Full model |  | Final model |  |
| --- | --- | --- | --- | --- | --- |
|  |  | Estimate | 95%-CI | Estimate | 95%-CI |
| Intercept | Constant | 0.9776*** | 0.8489, 1.1079 | 0.9233*** | 0.8623, 0.9894 |
| Group | Cases | 0.0708*** | 0.0330, 0.1086 | 0.0712*** | 0.0332, 0.1062 |
| Symptomatic | Month 6 | -0.0892*** | -0.1206, -0.0588 | -0.0919*** | -0.1228, -0.0623 |
| Age | numeric | -0.0008 | -0.0020, 0.0002 | -0.0009 | -0.0019, 0.0001 |
| Sex | Female | -0.0071 | -0.0316, 0.0198 | - | - |
| Pregnant | Yes | -0.0066 | -0.1629, 0.0757 | - | - |
| Ethnicity<sup>a</sup> | Asian/Asian British | Reference | Reference | - | - |
|  | Black / African/ Caribbean/ Black British | 0.0482 | -0.0089, 0.1059 | - | - |
|  | Other ethnic group | -0.084 | -0.2290, 0.0079 | - | - |
|  | White British/ Other White | -0.0413 | -0.0797, 0.0062 | - | - |
| Region | East Midlands | Reference | Reference | - | - |
|  | East of England | 0.0002 | -0.0763, 0.0754 | - | - |
|  | London | 0.045 | -0.0093, 0.1037 | - | - |
|  | North East | 0.029 | -0.0329, 0.0941 | - | - |
|  | North West | 0.0192 | -0.0452, 0.0860 | - | - |
|  | South East | 0.0329 | -0.0204, 0.0954 | - | - |
|  | South West | 0.0084 | -0.0440, 0.0770 | - | - |
|  | West Midlands | 0.0132 | -0.0545, 0.0743 | - | - |
|  | Yorkshire and The Humber | 0.0479 | -0.0012, 0.1126 | - | - |
| Comorbidities | None | 0.0362* | 0.0049, 0.0728 | 0.0382* | 0.0080, 0.0736 |
| Shielding | Yes | -0.0673* | -0.1346, -0.0133 | -0.0706** | -0.1407, -0.0182 |
| Household size | numeric | -0.0041* | -0.0127, 0.0020 | -0.0043** | -0.0128, 0.0020 |
| Vaccinated | Seasonal influenza (2020-2021) | -0.0185 | -0.0482, 0.0111 | -0.0191 | -0.0475, 0.0104 |
|  | COVID-19 (2 doses) | -0.0398 | -0.1276, 0.0742 | - | - |
| adj. R-squared |  | 0.095 |  | 0.093 |  |
| AIC |  | -385.2 |  | -407.2 |  |
| BIC |  | -278.5 |  | -365.3 |  |

Note: Negative values represent worse HRQoL and positive values represent improved HRQoL.
a: Answers of "Mixed/Multiple ethnicity" were combined with "Other ethnic group" given very small numbers. COVID-19 vaccination status not being significant in the final model may be impacted by the partial protection in vaccinated individuals, and our sample having been tested before the vaccination rollout started in England in December 2020.
Bias-corrected and accelerated (bca) confidence intervals (95%-CI) were based on the nonparametric bootstrap. Covariates with significance levels below $p < 0.1$ were entered in the multivariable analysis.
\*\*\* $p < 0.001$ , \*\* $p < 0.01$ , \* $p < 0.05$

## Discussion

This study explored the long-term health-related quality of life impact of non-hospitalised, laboratory-confirmed COVID-19 cases in England. Our cohort of cases experienced a lasting impact from physical symptoms, with the reported prevalence of symptoms similar to that reported in a systematic review of long COVID studies after 4 weeks and up to six months [1]. In addition, our study estimated substantial QALY losses for cases with persistent symptoms by month 6 that increased by age. Cumulatively, the total COVID-19-related QALY loss from morbidity in England by mid-September 2021 was also substantially higher than the COVID-19-related mortality in age groups younger than 45-54 years. This estimate may be conservative as we did not include the health burden from hospitalised, non-fatal cases [26], and because of under-reporting of non-fatal COVID-19 cases, particularly early on in the epidemic [27]. However, it stresses the growing importance of the COVID-19-related morbidity across age groups [25], and the shortcoming of using deaths and hospitalisations as the main metrics for the severity of an epidemic.

Furthermore, the cross-sectional analysis showed that cases without symptoms at month 6 had higher EQ-5D index utilities than controls without symptoms at month 6 (i.e., they were in better health), possibly due to the slightly improved levels of pain / discomfort and anxiety / depression (even though they were non-significant in the logistic regression). These results may point towards coping mechanisms in cases and potential mental health impacts felt from the COVID-19 pandemic and resultant restrictions (once the physical health is restored), as previously shown for the general populations in multiple countries [28-30]. Some of the HRQoL loss in controls may also come from isolation as those testing negative may still have been isolating until they were informed about the negative test result and reporting symptoms of other respiratory diseases. Other important variables to describe our samples were comorbidities, age and household size, but the predictive ability of the models may need to be interpreted with caution.

Previous studies on the QALYs lost in COVID-19 cases have been limited and focussed on the acute illness [31]. Compared to previous studies on the H1N1pdm2009 pandemic influenza virus, our estimates are higher than the estimated mean of 2.9 (range: 0-9.9) QALDs lost in laboratory-confirmed influenza cases in the UK [12] and of 3.3 (95% CI, 2.6–4.0) QALDs lost in Spain [11]. The differences may partly be explained by a more severe and/or longer-lasting health impact of COVID-19, and how the general population values their health differently during the COVID-19 pandemic [32].

### Strengths and Limitations

This study is one of the first to provide long-term QALY estimates associated with non-hospitalised COVID-19 in the UK, which will be of use in evaluation studies of vaccines, drugs, and non-pharmaceutical interventions. We also recruited a control group for the cross-sectional analysis, which is not often available in other studies.

As cases were recruited after testing positive for SARS-CoV-2, they completed versions of the EQ-5D three times in the initial survey, including retrospectively about their pre-COVID-19 baseline health. Such a design is common, and while this may introduce recall bias most cases in our sample reported symptom onset within 2 weeks of having been tested and thus completed the initial survey within 4 weeks. For controls, the survey was sent by email to reduce the administrative burden and costs, which we tried to compensate for by inviting a larger sample. For the cases, we found some indication that the retainment of participants with worse health may be higher in those who return surveys by post (which we accounted for in the models). Although all respondents were randomly sampled and we applied post-hoc weighting by age group and sex for more representative results, we also cannot rule out self-selection bias into this study by more health-conscious individuals.

We also provided results for both utility value sets in England of the EQ-5D-3L and EQ-5D-5L given that mapping from the responses on the EQ-5D-5L to the EQ-5D-3L is currently recommended by NICE [14, 15], but this position has been criticised [33].

Our drop in responses after the initial survey may partly be the result of responders’ fatigue, interference from the festive season in December, and cases who recovered and stopped returning follow-up surveys (as more than two-third of cases reported no issues at their last recorded observation). Our study was also only powered to detect a QALY loss in non-hospitalised cases. We did not calculate QALYs for children aged younger than 12 years in the absence of a validated EQ-5D value set, and the adult value sets likely being inadequate to be used in children [34]. Furthermore, we did not enquire about the impact on other family members and partners who have shown to be impacted, too [35].

In conclusion, our study quantified the QALY loss per case due to COVID-19 in England for a prolonged time after the acute illness episode, which increased by age and was higher for cases who reported symptoms at 6 months. We also showed that there is a substantial quality of life burden due to non-fatal COVID-19 at a population level, and particularly in younger ages.

## Data Availability

A waiver of ethical approval was received for this study from the PHE Research Support and Governance Office. Ethical approval for this study was not required as the collection of information on patients quality of life is part of the statutory tasks and routine surveillance activities of Public Health England and its legal successor, the UK Health Security Agency.

## Acknowledgements

We thank Archana N Purohit, Paul Charter, Charlotte Ryan, and Molly Viggars for administrative support.

## Technical appendix

### Details on study design and conduct

Surveys were sent out electronically using Snap Surveys (www.snapsurveys.com), and the initial survey was also sent by post to all participants to increase response rates. Subsequent surveys were sent out via the preferred method that the participant responded to the first survey. In follow-up postal surveys, we provided the option for respondents to add an email address if they wanted to switch to responding online. Follow-up reminders were sent by email for those that had not yet responded to the online survey.

### Details on data processing and cleaning

We cleaned the data by correcting inconsistencies (e.g. individuals reporting not being ill while also reporting symptoms like coughing) and data entry errors (e.g. calendar dates in the future). Some individuals returned responses to us via both the online survey and by post; for survey 1 when a completely filled-in online response was available we kept the online record (n=6), while for partially filled-in surveys online we manually entered the postal responses (n=3). For the follow-up surveys there were 16 duplicate responses by the same participants via both post and online (2.9%), and for diverging answers we considered the mean score on the VAS, duration of illness, and the date of the survey response, and otherwise we considered the highest value for the remaining fields (i.e., presence of symptoms coded as 1 vs not-present coded with 0, and for the EQ-5D responses higher values equal more limitations and worse health). One survey was returned by the spouse of a participant; given that we recruited participants matched by region but not by sex we thus kept the response. We also experienced a technical problem with survey 1 that resulted in the IDs of respondents not being captured. Hence respondents had to be manually matched against the cohort that responded to the survey using all the available demographic information and email addresses; despite best efforts 69 of the 566 (12.2%) non-duplicate responses to survey 1 were unable to be matched.

Furthermore, we calculated the time since individuals were tested based on the date of their response to each survey. This date was automatically recorded within the online survey and there was a field for the date that individuals completed the questions on the postal surveys. For postal surveys with a missing date field, we used the date that the response was received minus 2 days to account for potential delays from postal delivery. To minimise bias, we further removed 11 responses to survey 1, and 12 responses from the first three follow-up surveys, due to a long delay in receipt which overlapped with the timing of the subsequent follow-up survey. Similarly, we removed 15 cases who reported receiving secondary care, and cases who did not submit responses to the baseline survey.

### Details on the QALY estimation

In sensitivity analysis, we used the utility value sets of Ireland (which is valuing worse health states lower [17]), the Western preference pattern combining the values of Canada, England, Netherlands, and Spain (valuing worse health states higher [18]), the USA (valuing better health states lower [19]), and Germany (valuing better health states higher [20]).

For the initial survey, we estimated the utility between the time of filling in survey 1, the worst day since feeling ill or being tested, and the pre-COVID-19 baseline assuming an equal split under a trapezoidal rule. For responses with missing durations but respondents indicating still feeling unwell, we used as proxy the duration of the time of being tested until filling in the survey at week 0; for those reporting not feeling ill we used a duration of 1 day. For the follow-up surveys, we based the durations on the individual dates of when responses were received and responses of the EQ-5D reflecting that day.

When analysing the total individual patient-level QALY loss over 6 months as the dependent variable, the constant of the marginal model indicates the background QALY loss of cases over 6 months, while the indicator variable provides an estimate of the additive QALY loss for cases reporting symptoms by month 6. In a sensitivity analysis we stratified the data by the indicator variable to test for its robustness (with the resulting coefficients being near-identical to the marginal model).

### Additional results

**Supplementary Figure 1:**
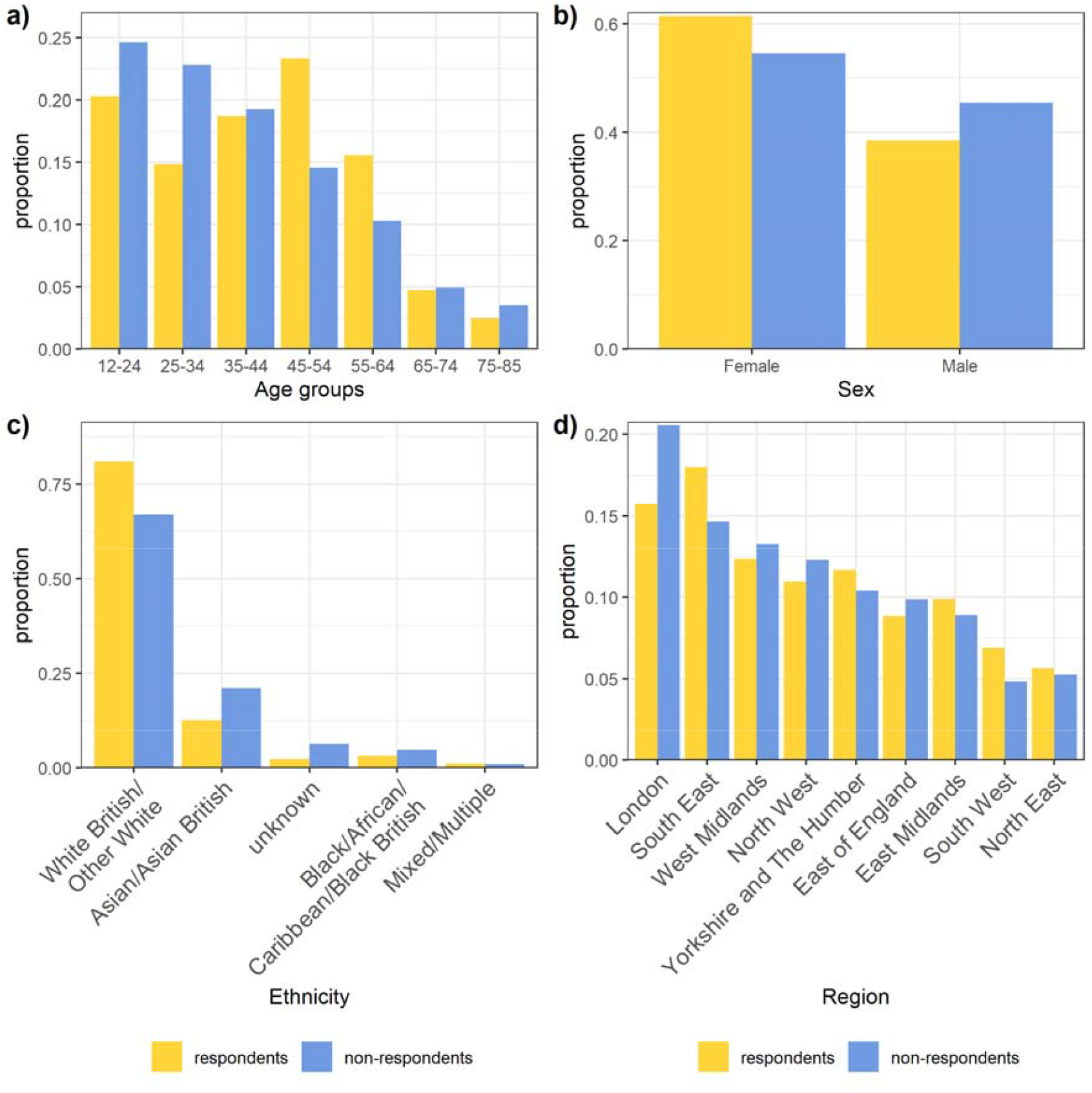
Comparison of the available demographic characteristics of the responding and non-responding cases (December 2020).

**Supplementary Figure 2:**
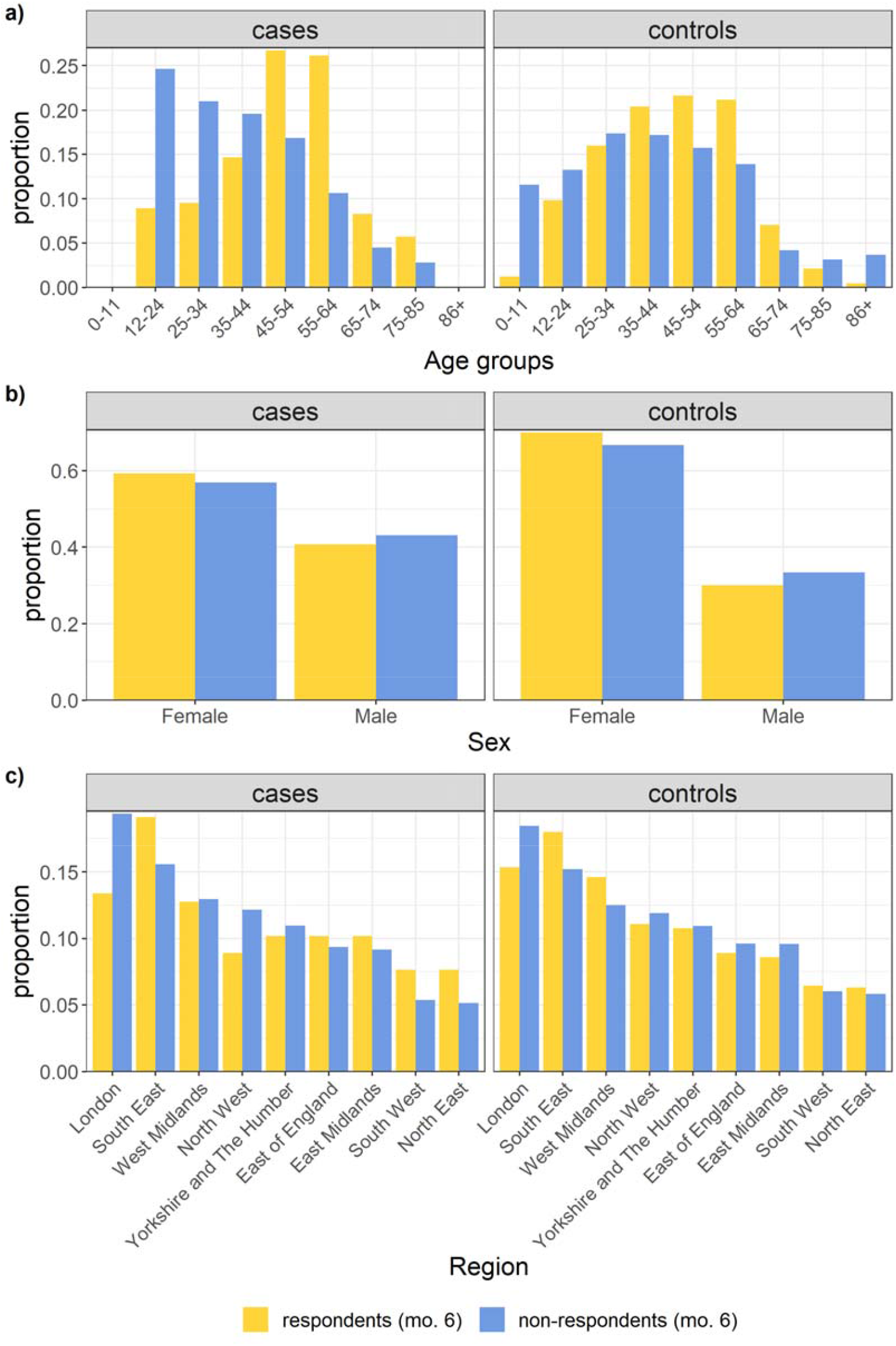
Comparison of the available demographic characteristics of the responding and non-responding cases and controls (June 2021). The results of the non-response include cases who were lost to follow up by June 2021. Ethnicity was not available for non-responding controls.

**Supplementary Figure 3:**
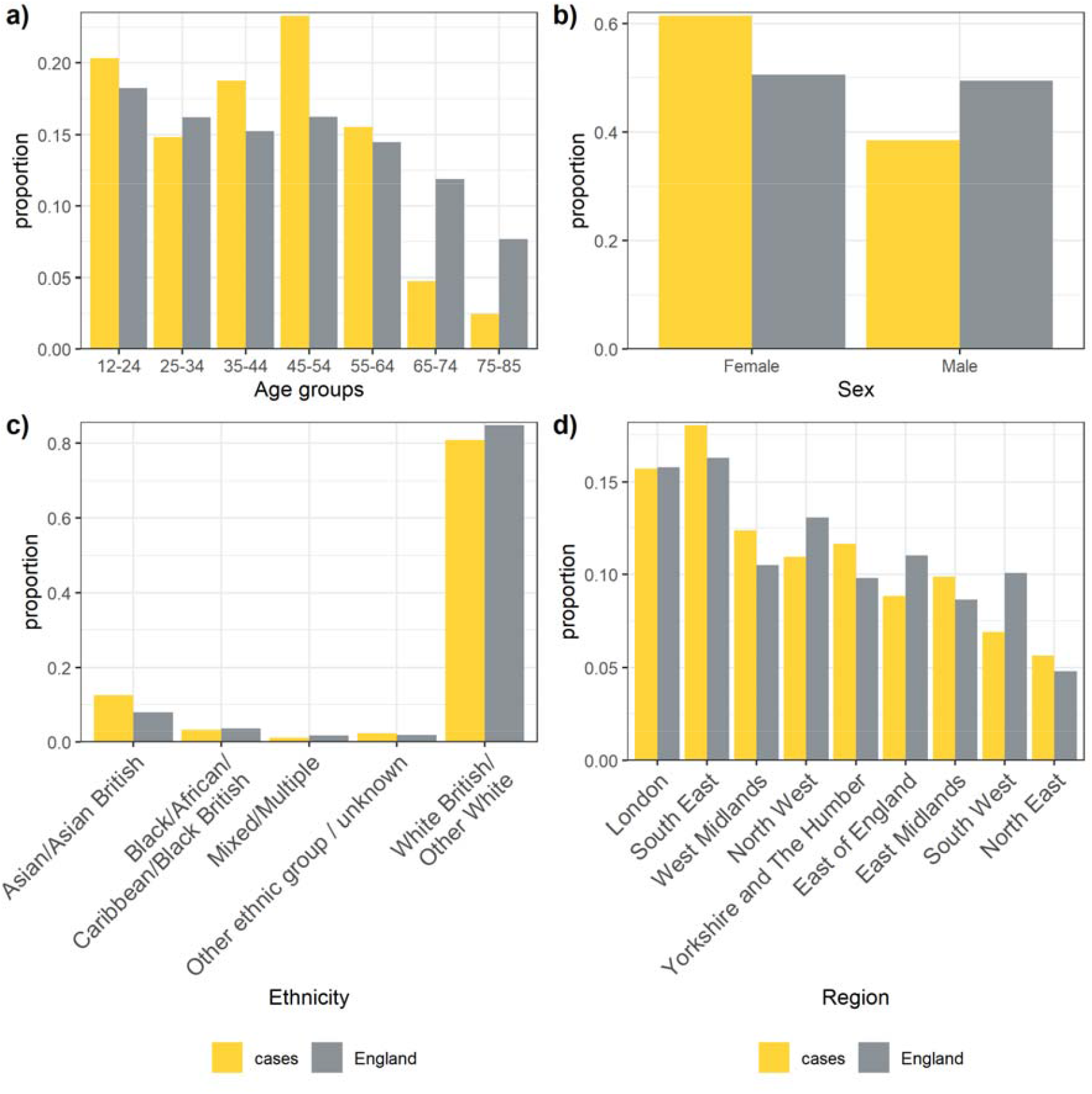
Comparison of the available demographic characteristics of the responding cases (December 2020) versus the population in England.

**Supplementary Figure 4:**
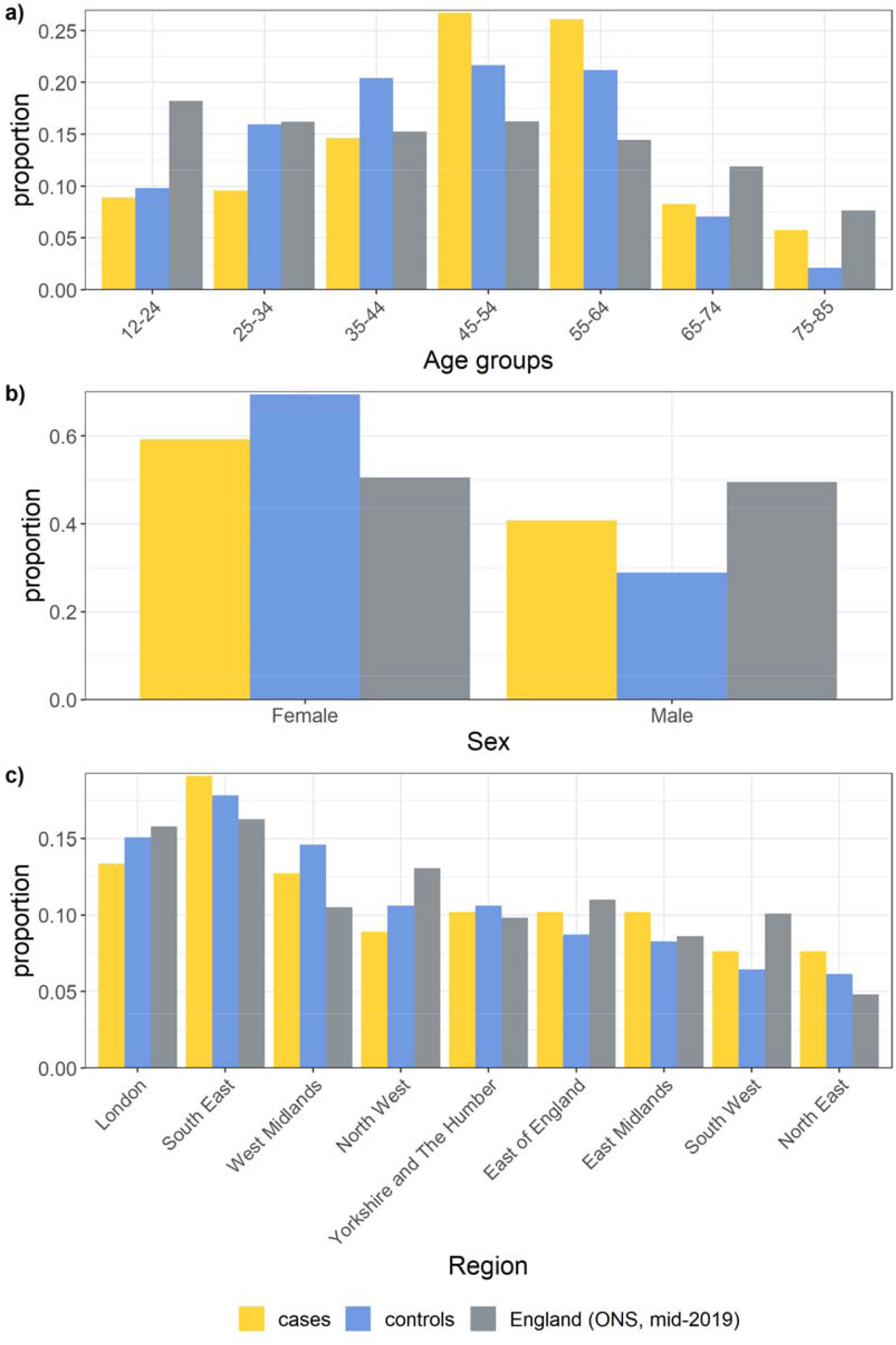
Comparison of the available demographic characteristics of the responding cases and controls (June 2021) versus the population in England.

**Supplementary Figure 5:**
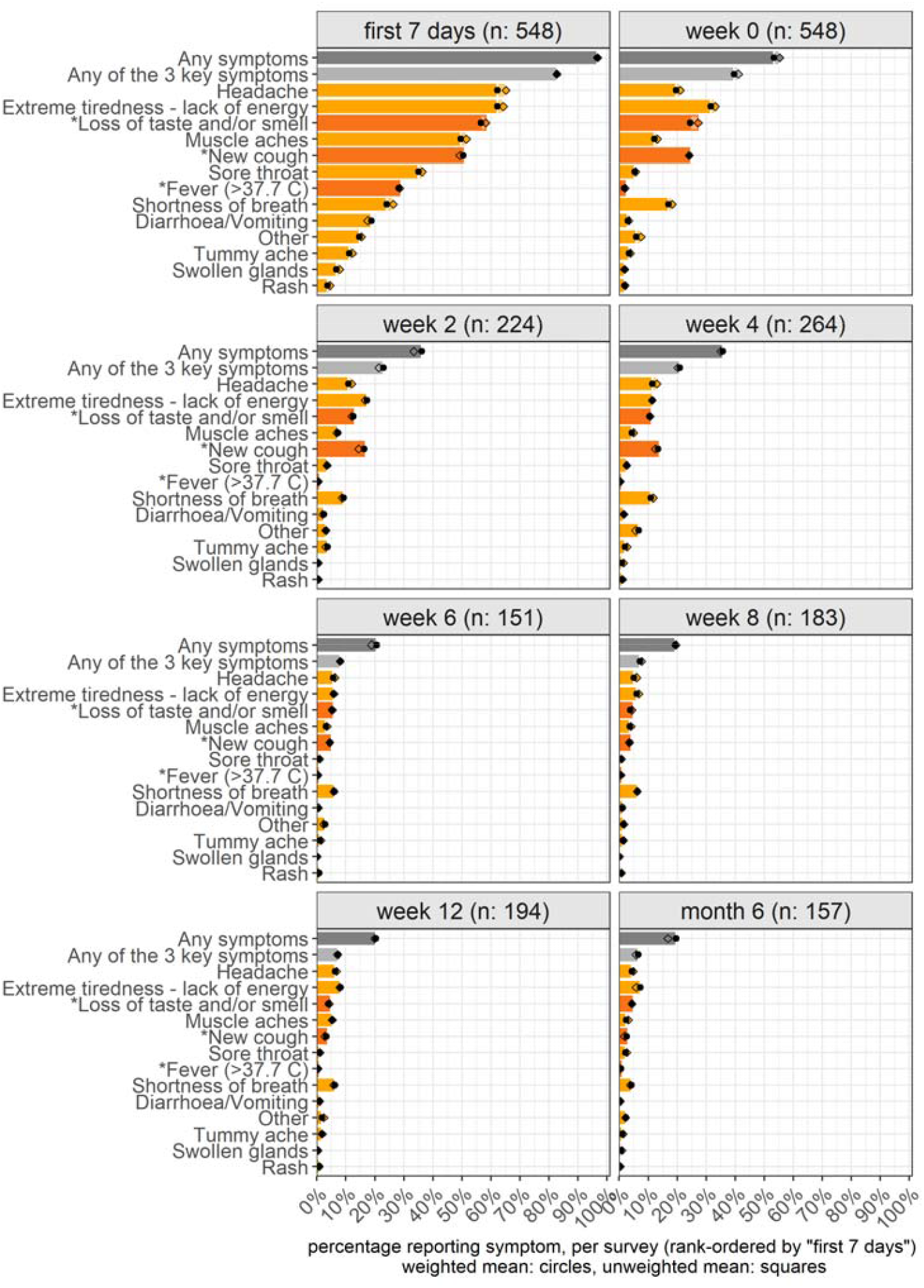
Physical symptoms reported in each survey by the SARS-CoV-2(+) cases over 6 months. Symptoms that have been seen as key indicators in the UK for members of the public to request a PCR test for COVID-19 are indicated in a darker colour and starred (i.e., fever of 38.8 C or higher, a new cough, or loss of taste and/or smell). The panels are rank-ordered by the proportion of symptoms in the first 7 days (top-left panel). Given the potential delay in symptom onset, getting PCR tested, and receiving the initial survey, the first survey enquired about the symptom status of cases within the first 7 days of feeling ill as well as if still feeling unwell today (week 0). Results are unweighted (squares) and weighted by age and sex of the population in England (filled circles).

**Supplementary Figure 6:**
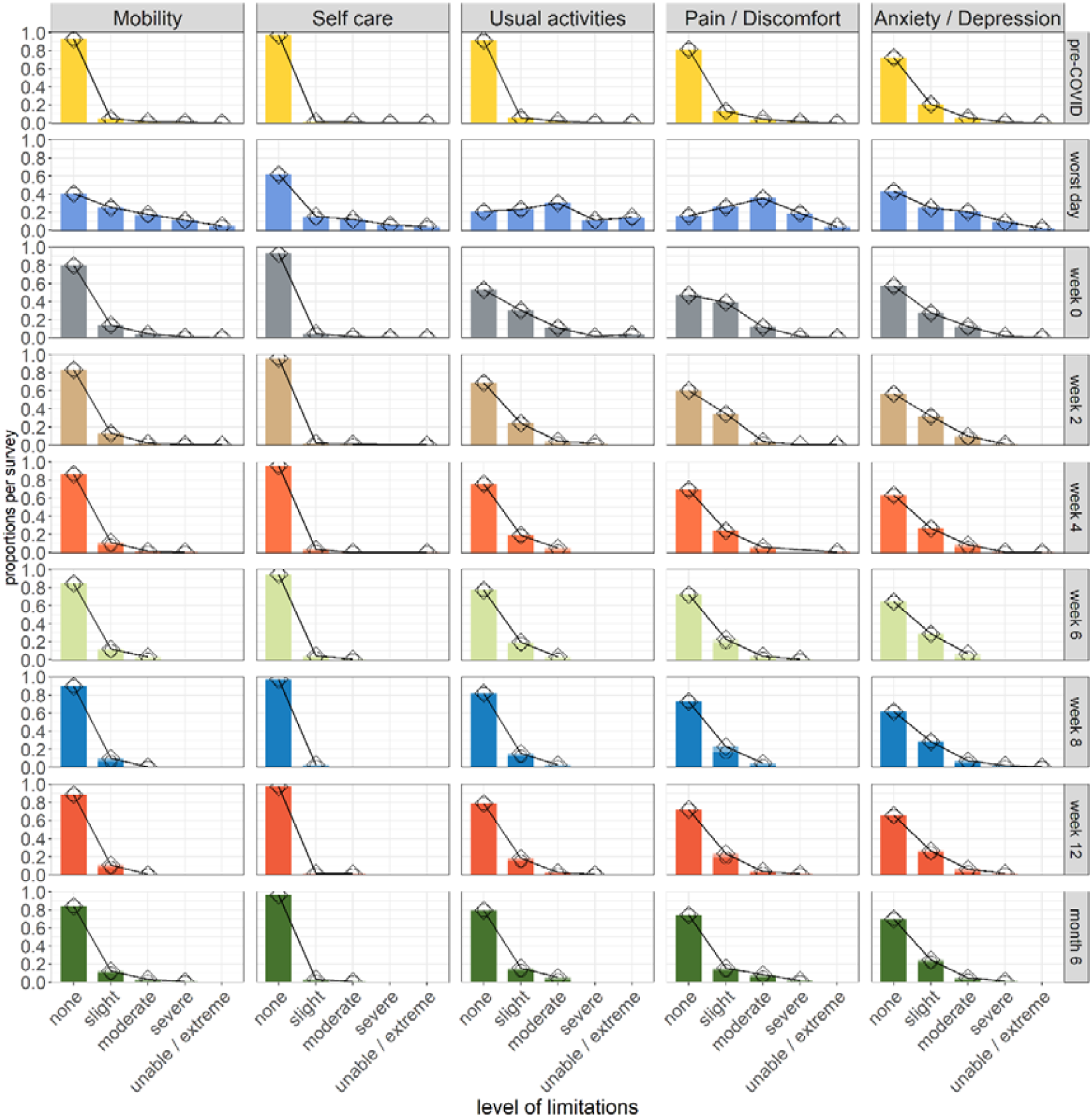
Frequency of responses on the EQ-5D-5L dimensions per survey. Results are unweighted (squares) and weighted by age and sex of the population in England (circles).

**Supplementary Figure 7:**
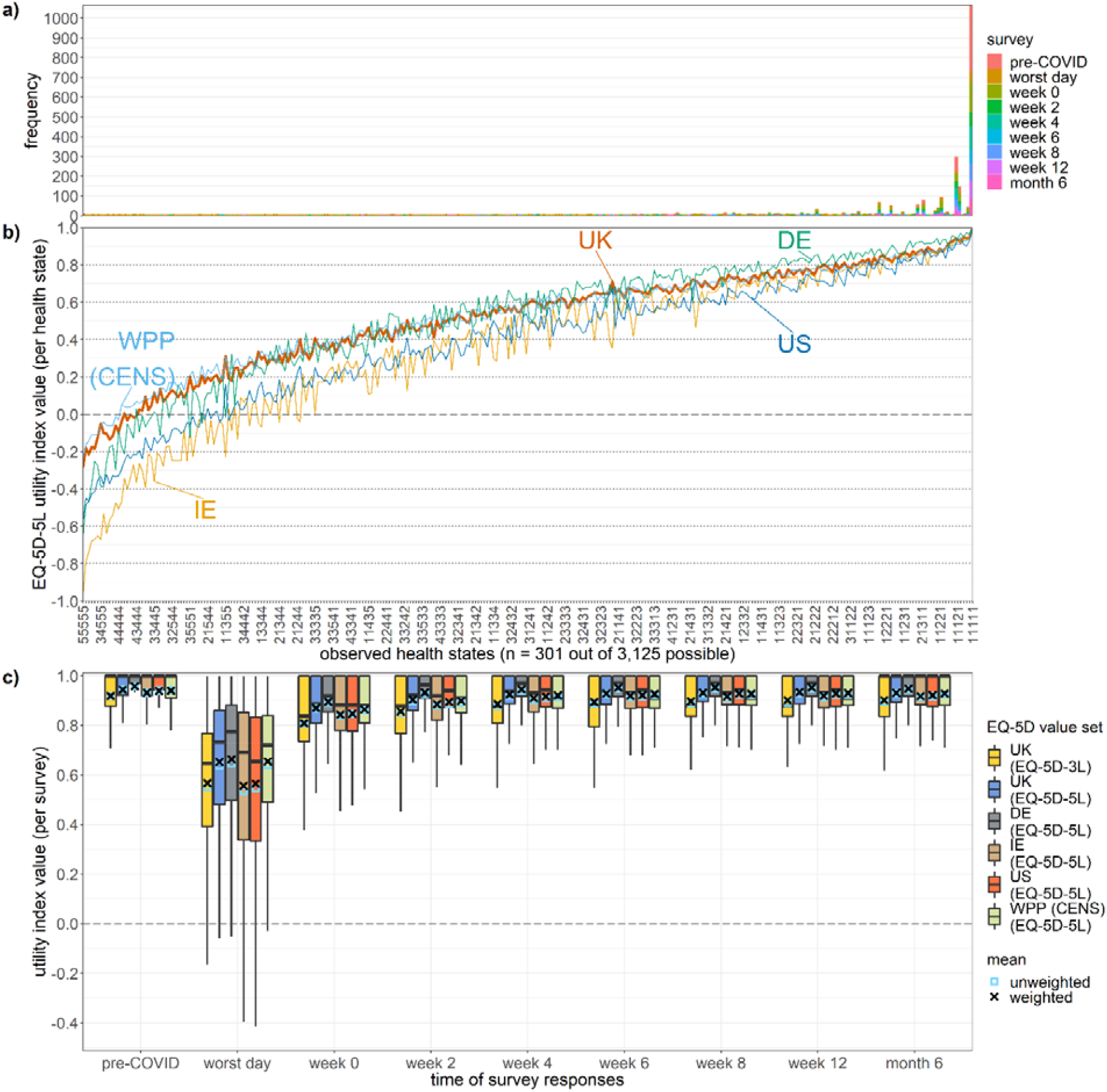
Frequency of health states reported by cases (panel a); utility index values for the reported health states as valued by different tariffs internationally (panel b); utility index scores per survey with different value sets (panel c). Given that the health states in our sample were leaning towards “better” health states with fewer limitations (i.e. towards “11111”; panel a), the EQ-5D value sets that assign lower index utility values to health states (from Ireland and the USA; panel b) resulted nonetheless in utility values per survey comparable to the other EQ-5D value sets (panel c). UK: United Kingdom; DE: Germany; IE: Ireland; US: United States of America; WPP (CENS): Western preference pattern of Canada, England, Netherlands and Spain.

**Supplementary Figure 8.**
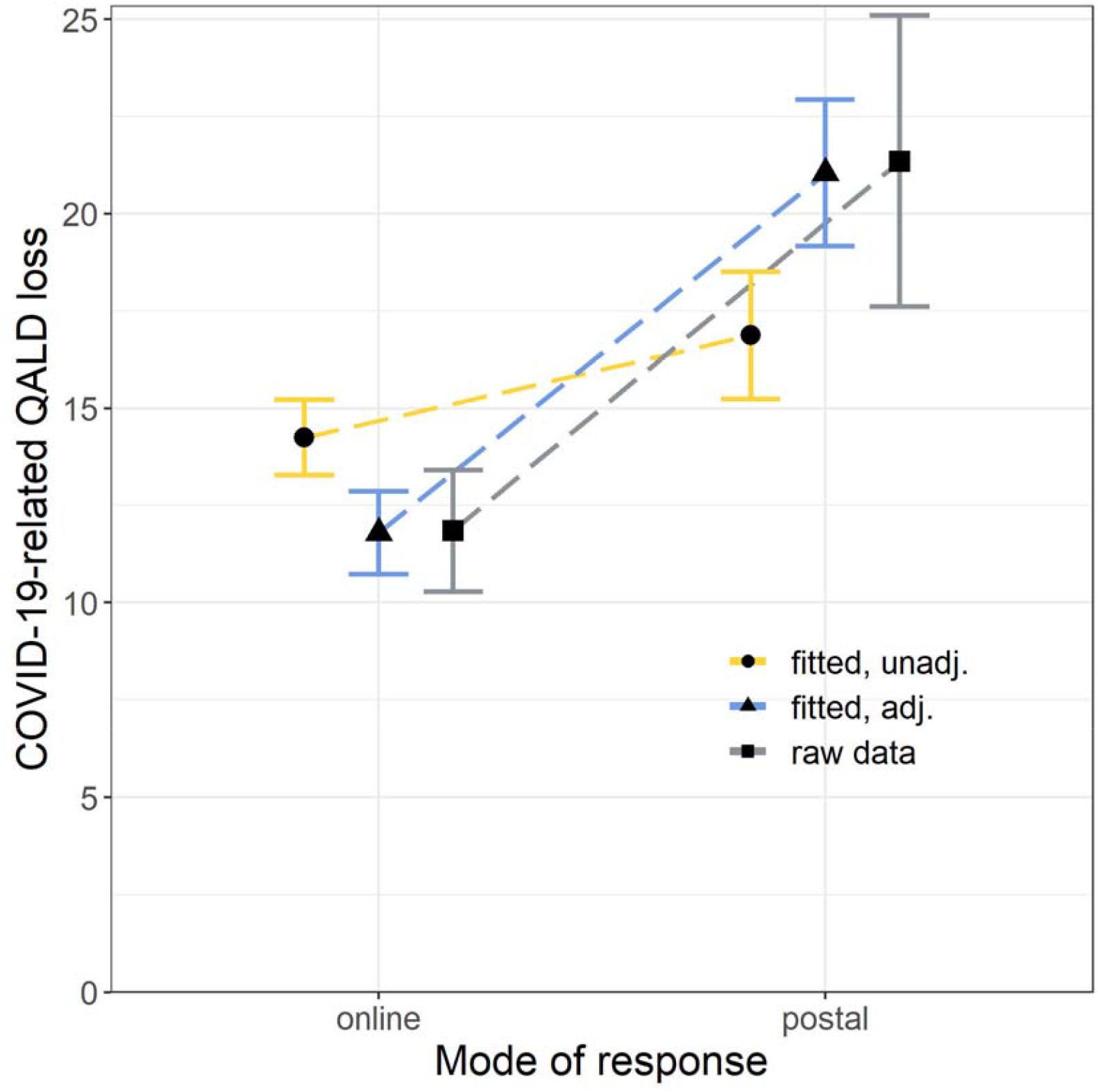
COVID-19-related QALD loss per case, separated by those who returned surveys online (n=357) versus those who preferred to return responses by post (n=191). The figure compares the raw data of the individual-patient QALY losses to the fitted results and provides some indication for the worse impact captured by the mode of response.

**Supplementary Table 1.**
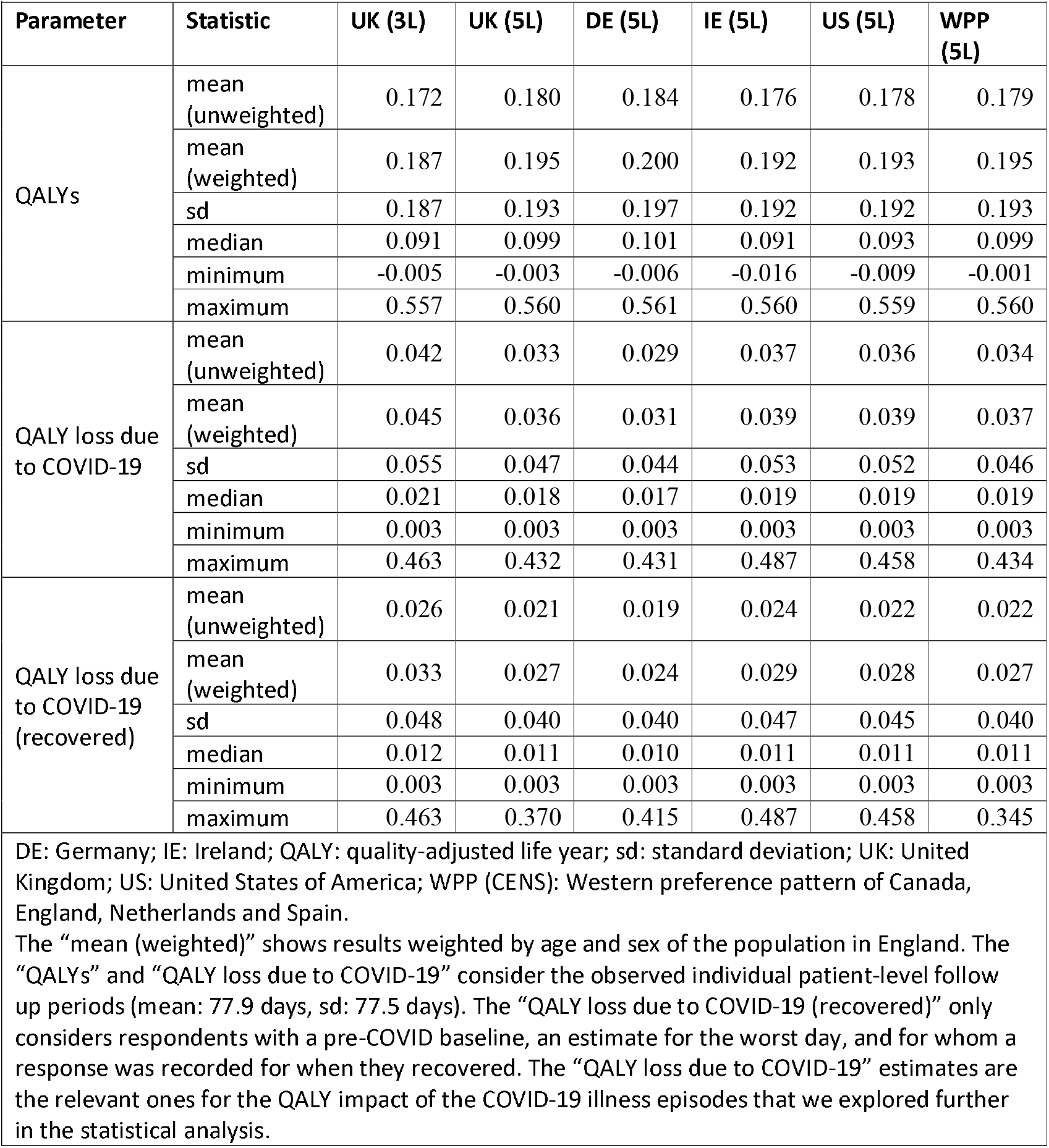
Unadjusted QALYs and QALY losses of the SARS-CoV-2(+) cases for different EQ-5D value sets.

**Supplementary Table 2:** Adjusted QALY losses of the SARS-CoV-2(+) cases, and SARS-CoV-2(+) cases who report symptoms at month 6.

|  |  |  |  |  |  |  |  |
| --- | --- | --- | --- | --- | --- | --- | --- |
| Value set (EQ-5D) | Variable | Marginal models (model 1) |  |  | Final models (model 2) |  |  |
|  |  | Estimate | Lower CI | Upper CI | Estimate | Lower CI | Upper CI |
| UK (3L) | Constant | 0.029 | 0.025 | 0.034 | - | - | - |
| UK (5L) |  | 0.024 | 0.021 | 0.028 | - | - | - |
| DE (5L) |  | 0.022 | 0.019 | 0.026 | - | - | - |
| IE (5L) |  | 0.027 | 0.024 | 0.032 | - | - | - |
| US (5L) |  | 0.026 | 0.022 | 0.030 | - | - | - |
| WPP (5L) |  | 0.025 | 0.022 | 0.029 | - | - | - |
| UK (3L) | Symptoms at month 6 | 0.075 | 0.060 | 0.090 | 0.061 | 0.045 | 0.076 |
| UK (5L) |  | 0.055 | 0.042 | 0.069 | 0.044 | 0.030 | 0.057 |
| DE (5L) |  | 0.043 | 0.032 | 0.059 | 0.033 | 0.020 | 0.046 |
| IE (5L) |  | 0.061 | 0.046 | 0.078 | 0.051 | 0.035 | 0.067 |
| US (5L) |  | 0.061 | 0.046 | 0.076 | 0.049 | 0.034 | 0.065 |
| WPP (5L) |  | 0.056 | 0.043 | 0.070 | 0.045 | 0.031 | 0.058 |
| CI: confidence interval, DE: Germany, IE: Ireland, UK: United Kingdom, US: United States, WPP: Western Preference Pattern (based on Canada, England, the Netherlands, and Spain).<br>Bias-corrected and accelerated 95%-CIs were based on the nonparametric bootstrap.<br>Model 1: Marginal models of reporting symptoms at month 6 against the QALY loss per EQ-5D value set.<br>Model 2: Final models of reporting symptoms at month 6 against the QALY loss per EQ-5D value set and adjusted for baseline utility, age, pregnancy, mode of response, having received the influenza vaccine in 2020-2021, and having received 2 COVID-19 vaccine doses (and the UK 3L, UK 5L, US, and WPP value set were additionally adjusted for the "household size"). |  |  |  |  |  |  |  |

**Supplementary Table 3:** Adjusted QALY losses of the SARS-CoV-2(+) cases, and by SARS-CoV-2(+) cases who report symptoms at month 6.

| Value set (EQ-5D) | Group | Unweighted results |  |  | Weighted results |  |  |
| --- | --- | --- | --- | --- | --- | --- | --- |
|  |  | Estimate | Lower 95%-CI | Upper 95%-CI | Estimate | Lower 95%-CI | Upper 95%-CI |
| UK (3L) | All cases | 0.041 | 0.038 | 0.044 | 0.044 | 0.033 | 0.054 |
| UK (5L) |  | 0.033 | 0.031 | 0.035 | 0.035 | 0.027 | 0.043 |
| DE (5L) |  | 0.029 | 0.027 | 0.030 | 0.030 | 0.024 | 0.037 |
| IE (5L) |  | 0.037 | 0.034 | 0.039 | 0.039 | 0.030 | 0.047 |
| US (5L) |  | 0.035 | 0.033 | 0.038 | 0.038 | 0.029 | 0.046 |
| WPP (5L) |  | 0.033 | 0.031 | 0.036 | 0.036 | 0.028 | 0.043 |
| UK (3L) | Cases without symptoms at month 6 | 0.029 | 0.027 | 0.031 | 0.030 | 0.023 | 0.037 |
| UK (5L) |  | 0.024 | 0.023 | 0.025 | 0.025 | 0.020 | 0.030 |
| DE (5L) |  | 0.022 | 0.021 | 0.023 | 0.023 | 0.018 | 0.027 |
| IE (5L) |  | 0.027 | 0.025 | 0.028 | 0.028 | 0.022 | 0.034 |
| US (5L) |  | 0.026 | 0.024 | 0.027 | 0.027 | 0.020 | 0.033 |
| WPP (5L) |  | 0.025 | 0.023 | 0.026 | 0.026 | 0.020 | 0.031 |
| UK (3L) | Cases with symptoms at month 6 | 0.101 | 0.097 | 0.105 | 0.101 | 0.080 | 0.107 |
| UK (5L) |  | 0.076 | 0.072 | 0.079 | 0.076 | 0.060 | 0.081 |
| DE (5L) |  | 0.062 | 0.059 | 0.065 | 0.062 | 0.048 | 0.067 |
| IE (5L) |  | 0.085 | 0.082 | 0.089 | 0.086 | 0.068 | 0.091 |
| US (5L) |  | 0.083 | 0.080 | 0.087 | 0.083 | 0.066 | 0.089 |
| WPP (5L) |  | 0.077 | 0.074 | 0.081 | 0.077 | 0.061 | 0.082 |

CI: confidence interval, DE: Germany, IE: Ireland, UK: United Kingdom, US: United States, WPP: Western Preference Pattern (based on Canada, England, the Netherlands, and Spain).
Results are based on the final regression models of the QALY loss per EQ-5D value set and adjusted for baseline utility, reporting symptoms at month 6, age, pregnancy, mode of response, having received the influenza vaccine in 2020-2021, and having received 2 COVID-19 vaccine doses (and the UK 3L, UK 5L, US, and WPP value set were additionally adjusted for the "household size"). We post-hoc weighted results based on age and sex of the population in England.

**Supplementary Table 4.** Multivariable linear regression models of the EQ-5D-5L utility index values of the SARS-CoV-2(+) cases and SARS-CoV-2(-) controls at month 6.

| Parameter | Category | Full model |  | Final model |  |
| --- | --- | --- | --- | --- | --- |
|  |  | Estimate | 95%-CI | Estimate | 95%-CI |
| Intercept | Constant | 0.9870*** | 0.8929, 1.0867 | 0.9322*** | 0.8846, 0.9815 |
| Group | Cases | 0.0524*** | 0.0222, 0.0834 | 0.0537*** | 0.0241, 0.0827 |
| Symptomatic | Month 6 | -0.0706*** | -0.0980, -0.0432 | -0.0719*** | -0.0983, -0.0462 |
| Age | numeric | -0.0005 | -0.0015, 0.0003 | -0.0005 | -0.0014, 0.0003 |
| Sex | Female | -0.0061 | -0.0253, 0.0166 | - | - |
| Pregnant | Yes | 0.0027 | -0.0933, 0.0608 | - | - |
| Ethnicity<sup>a</sup> | Asian/Asian British | Reference | Reference | - | - |
|  | Black / African/ Caribbean/ Black British | 0.0309 | -0.0085, 0.0763 | - | - |
|  | Other ethnic group | -0.0613 | -0.1836, 0.0062 | - | - |
|  | White British/ Other White | -0.034 | -0.0627, 0.0065 | - | - |
| Region | East Midlands | Reference | Reference | - | - |
|  | East of England | -0.0032 | -0.0633, 0.0535 | - | - |
|  | London | 0.0276 | -0.0113, 0.0720 | - | - |
|  | North East | 0.0181 | -0.0353, 0.0663 | - | - |
|  | North West | 0.0154 | -0.0321, 0.0719 | - | - |
|  | South East | 0.0253 | -0.0150, 0.0708 | - | - |
|  | South West | 0.007 | -0.0328, 0.0591 | - | - |
|  | West Midlands | 0.0037 | -0.0476, 0.0534 | - | - |
|  | Yorkshire and The Humber | 0.035 | -0.0053, 0.0796 | - | - |
| Comorbidities | None | 0.0332** | 0.0082, 0.0640 | 0.0341** | 0.0088, 0.0639 |
| Shielding | Yes | -0.0644** | -0.1324, -0.0134 | -0.0667** | -0.1390, -0.0164 |
| Household size | numeric | -0.002 | -0.0071, 0.0023 | - | - |
| Vaccinated | Seasonal influenza (2020-2021) | -0.0114 | -0.0344, 0.0124 | -0.0126 | -0.0340, 0.0110 |
|  | COVID-19 (2 doses) | -0.0312 | -0.0978, 0.0531 | - | - |
| adj. R-squared |  | 0.088 |  | 0.088 |  |
| AIC |  | -721.2 |  | -749.4 |  |
| BIC |  | -614.4 |  | -712.2 |  |

a: Answers of “Mixed/Multiple ethnicity” were combined with “Other ethnic group” given very small numbers. COVID-19 vaccination status not being significant in the final model may be impacted by the partial protection in vaccinated individuals.
Bias-corrected and accelerated (bca) confidence intervals (95%-CI) were based on the nonparametric bootstrap. Covariates with significance levels below $p < 0.1$ were entered in the multivariable analysis.
\*\*\* $p < 0.001$ , \*\* $p < 0.01$ , \* $p < 0.05$

